# Bridging the Literacy Gap for Surgical Consents: An AI-Human Expert Collaborative Approach

**DOI:** 10.1101/2023.05.06.23289615

**Authors:** Rohaid Ali, Ian D. Connolly, Oliver Y. Tang, Fatima N. Mirza, Benjamin Johnston, Hael A. Abdulrazeq, Paul F. Galamaga, Tiffany J Libby, Neel R. Sodha, Michael W. Groff, Ziya L. Gokaslan, Albert E. Telfeian, John H. Shin, Wael F. Asaad, James Zou, Curtis E. Doberstein

**Author notes:** Corresponding Author: Rohaid Ali, MD. Department of Neurosurgery, Brown University/ Rhode Island Hospital, Providence, RI 02903. Mailing Address: LPG Neurosurgery, 593 Eddy Street, APC6, Providence, RI 02903. Drs Ali, Connolly, and Mr. Tang contributed equally to this work. **Conflicts of Interest:** The authors report no conflict of interest concerning the materials or methods used in this study or the findings specified in this paper. **Disclosure of Funding:** The authors have no funding relevant to the conduct of this study to disclose. **Details of Previous Presentation:** None.

## Abstract

**Background:** Despite the importance of informed consent in healthcare, the readability and specificity of consent forms often impedes patients’ comprehension. Health literacy is linked to patient outcomes, making it essential to address these issues. This study investigates the use of GPT-4 to simplify surgical consent forms and introduces an AI-human expert collaborative approach to validate content appropriateness.

**Methods:** Consent forms from multiple institutions were assessed for readability and simplified using GPT-4, with pre- and post-simplification readability metrics compared using nonparametric tests. Independent reviews by medical authors and a malpractice defense attorney were conducted. Finally, GPT-4’s potential for generating *de novo* procedure-specific consent forms was assessed, with forms evaluated using a validated 8-item rubric and expert subspecialty surgeon review.

**Results:** Analysis of 15 academic medical centers’ consent forms revealed significant reductions in average reading time, word rarity, and passive sentence frequency (all *P*<0.05) following GPT-4-faciliated simplification. Readability improved from an average college freshman to an 8th-grade level (*P*=0.004), matching the average American’s reading level. Medical and legal sufficiency consistency was confirmed. GPT-4 generated procedure-specific consent forms for five varied surgical procedures at an average 6th-grade reading level. These forms received perfect scores on a standardized consent form rubric and withstood scrutiny upon expert subspeciality surgeon review.

**Conclusions:** This study demonstrates the first AI-human expert collaboration to enhance surgical consent forms, significantly improving readability without sacrificing clinical detail. Our framework could be extended to other patient communication materials, emphasizing clear communication and mitigating disparities related to health literacy barriers. Ensuring AI technologies are safely incorporated into clinical practice is crucial to reach a wide range of patients, including the most vulnerable.

**Data Availability Statement:** The raw data was not publicly deposited due to incorporating proprietary datasets, such as the Corpus of Contemporary American English. However, data are available on request for replicability.

**Code Availability Statement:** The code used for these analyses are available from the authors on request.

## Introduction

Informed consent is a fundamental ethical principle and legal requirement in health care, ensuring that patients are provided with adequate information to make informed decisions about their treatment options. Consent forms serve as both an educational tool and legal documentation of the discussion that protects both patient and physician. When well-designed, the surgical consent provides patients with clear, concise, and comprehensible information regarding the risks, benefits, and alternatives of the proposed surgical procedure. However, a significant challenge in achieving truly informed consent lies in the readability of these forms.^1–3^ Previous research has shown that a substantial proportion of surgical consent forms are written at a reading level that exceeds the average patient’s comprehension.^1–4^ While most consent forms are written at a college freshman level or higher, the average reading level of American adults is equivalent to that of an 8^th^ grade student.^1–4^ This discrepancy is especially relevant in light of extensive research demonstrating that health literacy is associated with patient outcomes due to its impact on factors such as care-seeking, health promotion behaviors, adherence to physician recommendations, and self-advocacy in a clinical setting.^5–8^ Accordingly, to optimize comprehension and decision-making in the perioperative environment, clinicians must reduce potential barriers to patient health literacy.

In addition to the readability challenge, procedural consent forms are often written in a “one-size-fits-all” format that is generalizable to any potential procedure but that fails to discuss procedure-specific characteristics, such as steps, risks, and benefits, with sufficient nuance. Surgeons may circumvent the problem of generic consent forms by providing verbal supplementation or by utilizing unique procedure-specific patient education literature or consents. However, the implementation of these measures faces several potential barriers, such as institutional requirements to maintain generic consents or constantly evolving evidence regarding the individualized risks and benefits of specific procedures, which necessitates continual expert review and scrutiny.

Navigating the twin challenges of administering consent forms that are too difficult to read or nonspecifically tailored to a specific procedure is difficult, as potential quality improvement solutions may require substantial investment in terms of time, resources, and human capital. Moreover, it is unclear if a modified consent that uses more straightforward language may compromise the thoroughness of the original form from a medicolegal standpoint. In light of such barriers, we propose a novel AI-human expert framework to address these problems. Newly-developed artificial intelligence systems, specifically in the form of large language models (LLMs), have shown promise in their ability to summarize, adjust, and re- formulate text in a manner that could be highly relevant to this task.^9, 10^ This study **a)** quantitatively and qualitatively investigates the application of the GPT-4 (OpenAI; San Francisco, CA) general LLM to assess and transform surgical consent forms into a more accessible reading level in an efficient, standardized, and effective manner; **b)** develops a streamlined and extensible framework involving medical and legal review to ensure that content remains the same between original and simplified consents, and **c)** verifies the ability of GPT-4 to generate highly readable and procedure-specific consents *de novo* that meet expert-level scrutiny. This approach has the potential to significantly enhance the consent process by providing patients with clear, comprehensible, and specific information, ultimately promoting truly informed consent.

## Methods

Consent forms were obtained from the authors’ own institutions and, for other institutions, publicly available consent forms were identified on their respective websites. These consents were “generic” in the sense that they did not refer to any particular procedure or operation. Based on common estimates, a reading speed of 200 words-per-minute was used to calculate total reading time for forms.^11^ To quantify the readability of the consent forms, we utilized Flesch-Kincaid Reading Level, Flesch Reading Ease score, and word rarity measurements. Flesch-Kincaid Reading Level is a validated and widely-used metric that measures readability in terms of United States grade level of education, based on average words per sentence and average syllables per word.^1–3^ The Flesch Reading Ease score uses these same two variables to calculate a score ranging from 0-100, with higher scores denoting improved readability. A score of ≥60 corresponds to below a 9^th^ grade reading level, whereas a score of ≤30 denotes a college graduate reading level.^3^ These readability scores were calculated automatically via Microsoft Word (Microsoft; Redmond, WA). Mean word rarity was calculated for each form by averaging the rarity of each lemma or root word (ex. operate is the lemma for “operated” and “operating”) in the overall text, following the exclusion of “stop words” encapsulating commonly used words, such as conjunctions and determiners.^12^ The *koRpus* package in R Version 4.1.2 (Foundation for Statistical Computing, Vienna, Austria) and TreeTagger software were used to divide text into semantic parts, tag part-of-speech, and merge lemmas with frequency data on the 61,000 most common lemmas calculated using the one billion word Corpus of Contemporary American English (COCA).^13^

Subsequently, we applied the GPT-4 model on April 2, 2023 to simplify each consent form by providing the following prompt: “While still preserving the same content and meaning, please convert to the average American reading level:” followed by the consent text. We then re-evaluated the readability measures post-simplification (**Figure 1**). Three medical authors (RA, IDC and HA) and one malpractice defense attorney author (PG) independently reviewed the consent forms before and after the conversion to ensure that the content remained comparable. Paired nonparametric Mann-Whitney tests were used to compare pre- and post-simplification readability metrics, with statistical significance assessed at *P*<0.05.

**Figure 1:**
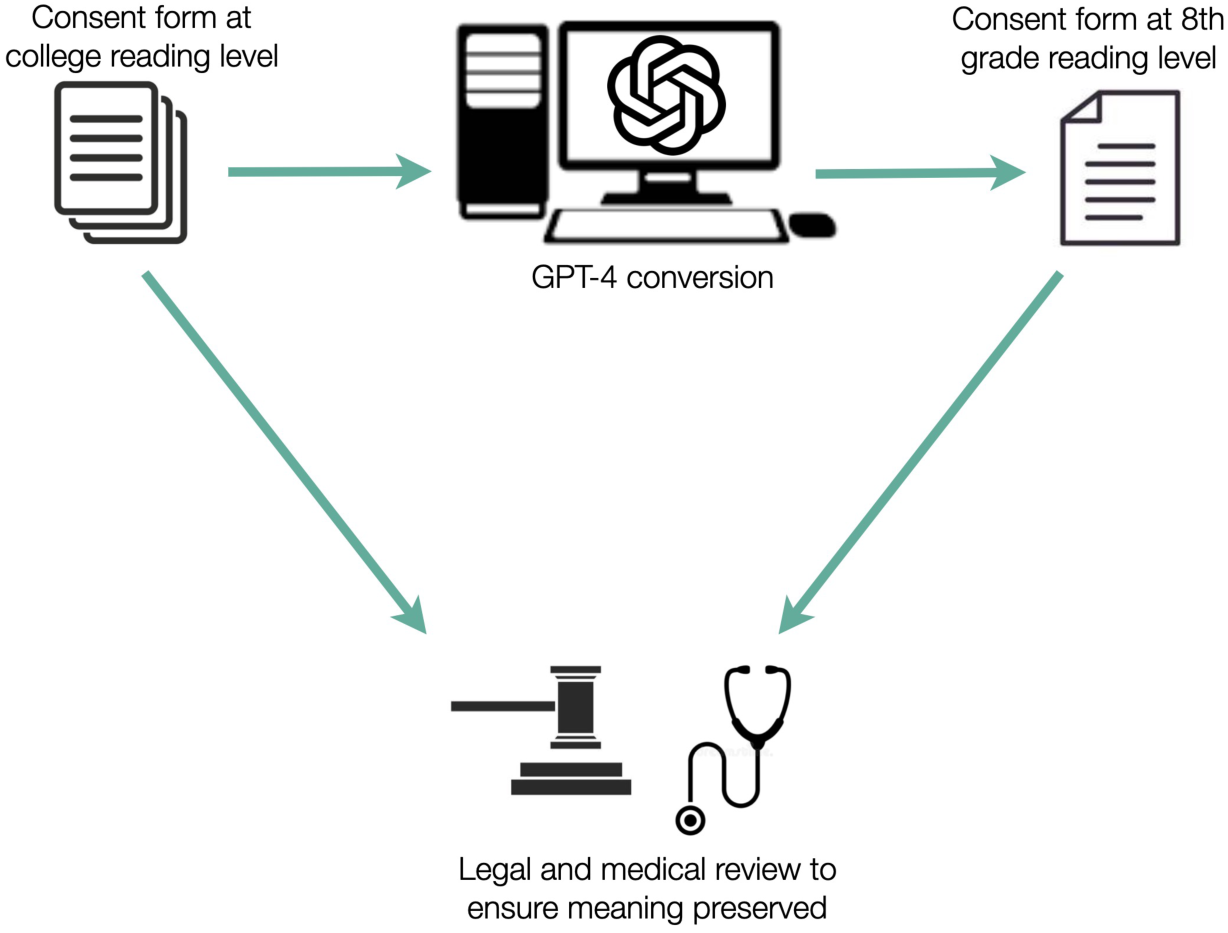
Methodology for Surgical Consent Form Readability Simplification. Schematic of methodology for surgical consent form readability simplification. A standardized prompt on GPT-4 was used to mediate simplification, with medical and legal review used to validate that meaning and quality was preserved.

In addition to the aforementioned methods for generic consents, we sought to explore the potential of GPT-4 for generating procedure-specific consent forms. We prompted GPT-4 on April 23, 2023 to create *de novo* surgical consent forms for five unique operations across the following surgical subspecialities: (1) cranial neurosurgery, (2) spine surgery, (3) general surgery, (4) cardiothoracic surgery, and (5) Mohs micrographic (i.e., dermatologic) surgery. The five chosen operations embody the diversity of surgical procedures in terms of locations (acute care, ambulatory, or clinic-based), acuity levels (emergent to elective), invasiveness, organ systems, and the variety of tools used in the surgical process (see Results for list of operations). Furthermore, we asked GPT-4 to produce text at the average American reading level. Subsequently, per the AI-human expert framework presented in this study, we utilized two independent arms of review to ensure the comprehensiveness and accuracy of *de novo* forms: procedure-specific expert review by subspecialty surgeons and a validated objective rubric for quantifying consent form quality.^14^ For the latter, we utilized an eight-item score ranging from 0-20 developed by Spatz e*t al.* for defining rigorous consent form requirements, incorporating input from Centers for Medicare and Medicaid Services, patients, and patient advocates. Items tracked by the score include clear language describing the procedure itself, benefits, quantitative and qualitative probability of risks, and alternatives. This score has been validated with high inter-rater agreement and used previously to assess inter-hospital variation in consent form quality.^14, 15^

## Results

Consent forms from 15 large academic medical centers were selected (**Supplementary Figure 1)**. By region, 7 were in the Northeast, 2 in the Mid-Atlantic, and 1 in each of the Southeast, South, Southwest, West, Northwest, and Midwest. 6 were publicly owned institutions (or their academic affiliation was tied to a public medical school), whereas the other 9 were private. All were affiliated with a medical school and serve as a teaching hospital. The average number of beds at each institution was 791 with a standard deviation of 256. All had Level 1 trauma center certification. 14 of the 15 institutions were primarily adult hospitals. Consent forms consisted of a median of 3,976.0 characters (interquartile range [IQR]=2,113.0–4,485.5 characters) and 651.0 words (IQR=396.0–803.0 words), requiring on average approximately 3 minutes and 15 seconds of reading time.

Following LLM-facilitated simplification of form text, three physician authors (RA, HA, and IDC) independently reviewed the consent forms before and after simplification to ensure that the content remained comparable, and all three agreed that the content remained comparable for all 15 consent forms. Additionally, one medical malpractice defense attorney (PG) reviewed the consents before and after simplification and determined that all 15 consent pairs met the same legal sufficiency.

After consent processing by the LLM, there was a significant decrease in median number of characters (before=3,967.0 vs. after=2,485.0 characters, *P*<0.001) and words (before=651.0 vs. after=483.0 words, *P*<0.001), decreasing reading time from 3.26 to 2.42 minutes (*P*<0.001, **Figure 2A**). Moreover, the median characters per word (before=5.4 vs. after=4.5 characters, *P*<0.001) and words per sentence (before=21.6 vs. after=19.0 words, *P*=0.001) decreased (**Figure 2**). There was additionally a significant decrease in the percentage of sentences written in the passive voice (before=38.4% vs. after=20.0%, *P*=0.024).

**Figure 2:**
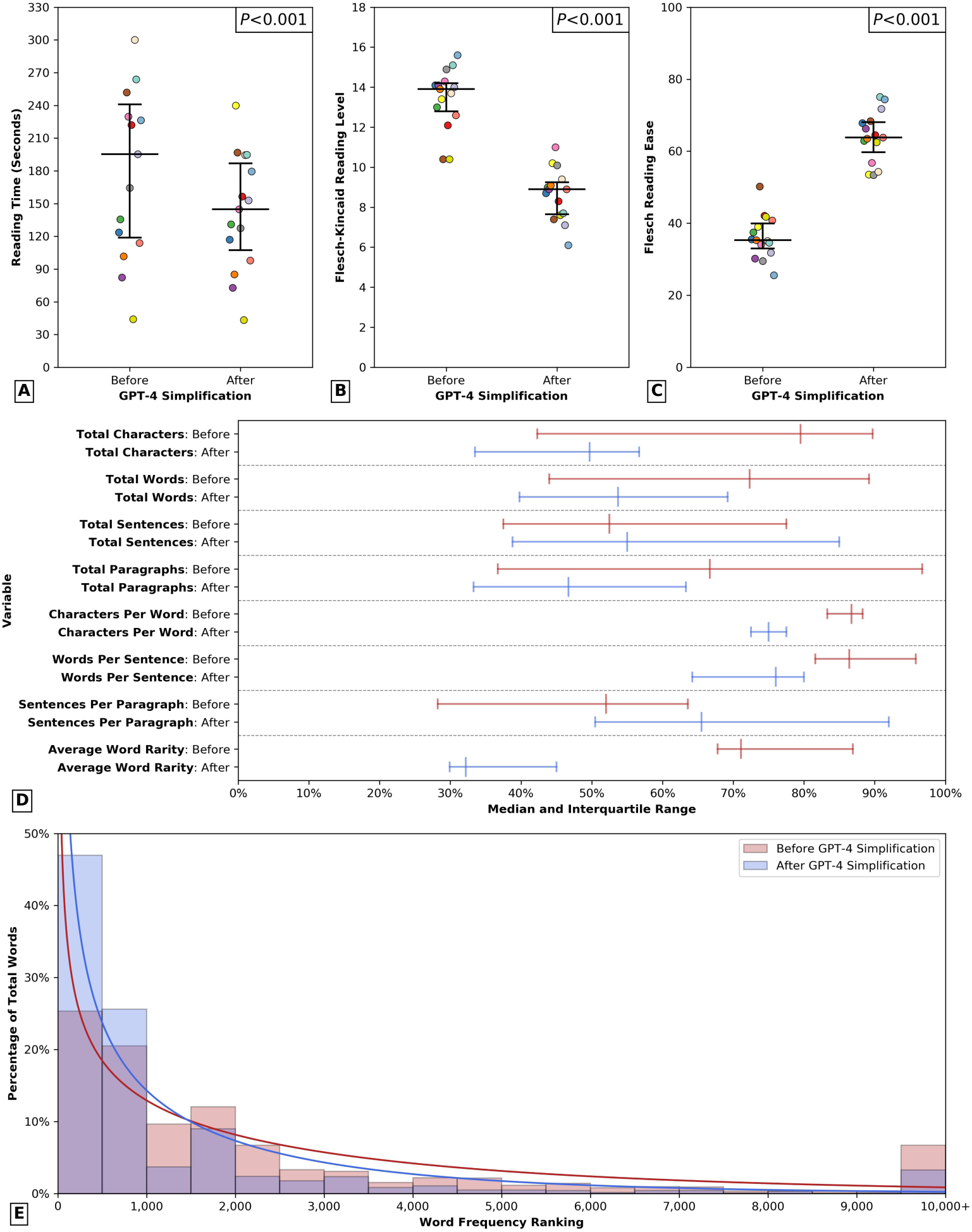
Differences in Surgical Consent Form Readability and Linguistic Parameters Before and After Simplification. Differences in surgical consent form readability before and after simplification mediated by GPT-4. Median and interquartile range for each variable are plotted. P-Values reported correspond to results from nonparametric Mann-Whitney tests. For panels A-C, a distinct color was used to label each individual institution (n=15). A: Differences in reading time. B: Differences in Flesch-Kincaid Reading Level. C: Differences in Flesch Reading Ease score. D: Differences in other linguistic variables before (red) and after (blue) simplification. Due to differences in scale between variables, all results were reported as a percentages (0-100%) of a predetermined constant: 5,000 for total characters, 900 for total words, 40 for total sentences, 30 for total paragraphs, 6 for characters per word, 25 for words per sentence, 2.5 for sentences per paragraph, and 4,000 for average word rarity. E: Histogram visualizing changes in distribution of word frequency ranking of consent form text before (blue) and after (red) simplification mediated by GPT-4. Word frequency in terms of rank within the English language is plotted on the x-axis, with higher rank denoting increased rarity and ranks 10,000 and above combined into a single category. Solid lines denote distribution of word frequency ranking following fitting the data to a γ distribution.

Prior to LLM processing, surgical consent forms had a median Flesch-Kincaid Reading Level of 13.9 (IQR=12.8–14.2; **Figure 2B**) and Flesch Reading Ease score of 35.3 (IQR=33.0– 39.9; **Figure 2C**), both denoting a college education level of readability difficulty. After simplification, there was a significant decrease in the Reading Level (before=13.9 vs. after=8.9, *P*=0.004) and improvement in Reading Ease (before=35.3 vs. after=63.8 *P*<0.001) from the reading level of a college freshman to that of an 8^th^ grade student. Moreover, the average rarity of the words used in the text decreased significantly after simplification (before=2,845 vs. after=1,328, *P*<0.001; **Figure 2E**).

Finally, for evaluation of procedure-specific consent forms, we prompted GPT-4 to generate consent forms for five diverse surgical procedures at the average American reading level and investigated if these generated consents aligned with the recommendations provided by Spatz *et al*.^14^ The procedure-specific consents included (**Figure 3A-E, all at end of manuscript**): (1) awake, subthalamic nucleus deep brain stimulating (STN-DBS) electrode placement with microelectrode recordings and test-stimulation for Parkinson’s disease (**Figure 3A**); (2) lumbar 4-5 (L4-5) percutaneous endoscopic lumbar discectomy (PELD) for L4 radiculopathy (**Figure 3B**,); (3) laparoscopic appendectomy for acute appendicitis (**Figure 3C**); (4) coronary artery bypass grafting (CABG) for acute Non-ST-segment Elevation Myocardial Infarction (NSTEMI) with multi-vessel coronary artery disease (**Figure 3D**; and (5) Mohs micrographic surgery for basal cell carcinoma (**Figure 3E**). The average word count of these procedure-specific forms was 414, with an estimated reading time of 2 minutes and 4 seconds. The Flesch-Kincaid Grade level was calculated to be 6.7, which corresponds to approximately the 6th grade in an American school. Three medical authors (RA, IDC, and HA) evaluated the generated consent forms using the rubric laid out by Spatz *et al.* and found that all five procedure-specific consents scored a perfect 20 out of 20.^14^ In other words, all consent forms met the minimum requirements, which included describing the procedure, explaining how the procedure will be performed, providing the clinical rationale, outlining patient-oriented benefits, presenting alternatives, and including a space to mark the date the consent form was signed. Despite the variety of procedures encapsulated by this task, procedure-specific expert review by subspecialty surgeons (WFA, AET, NRS, NRS, and TJL, respectively) yielded no wording changes or significant clinical inaccuracies requiring correction.

**Figure 3:**
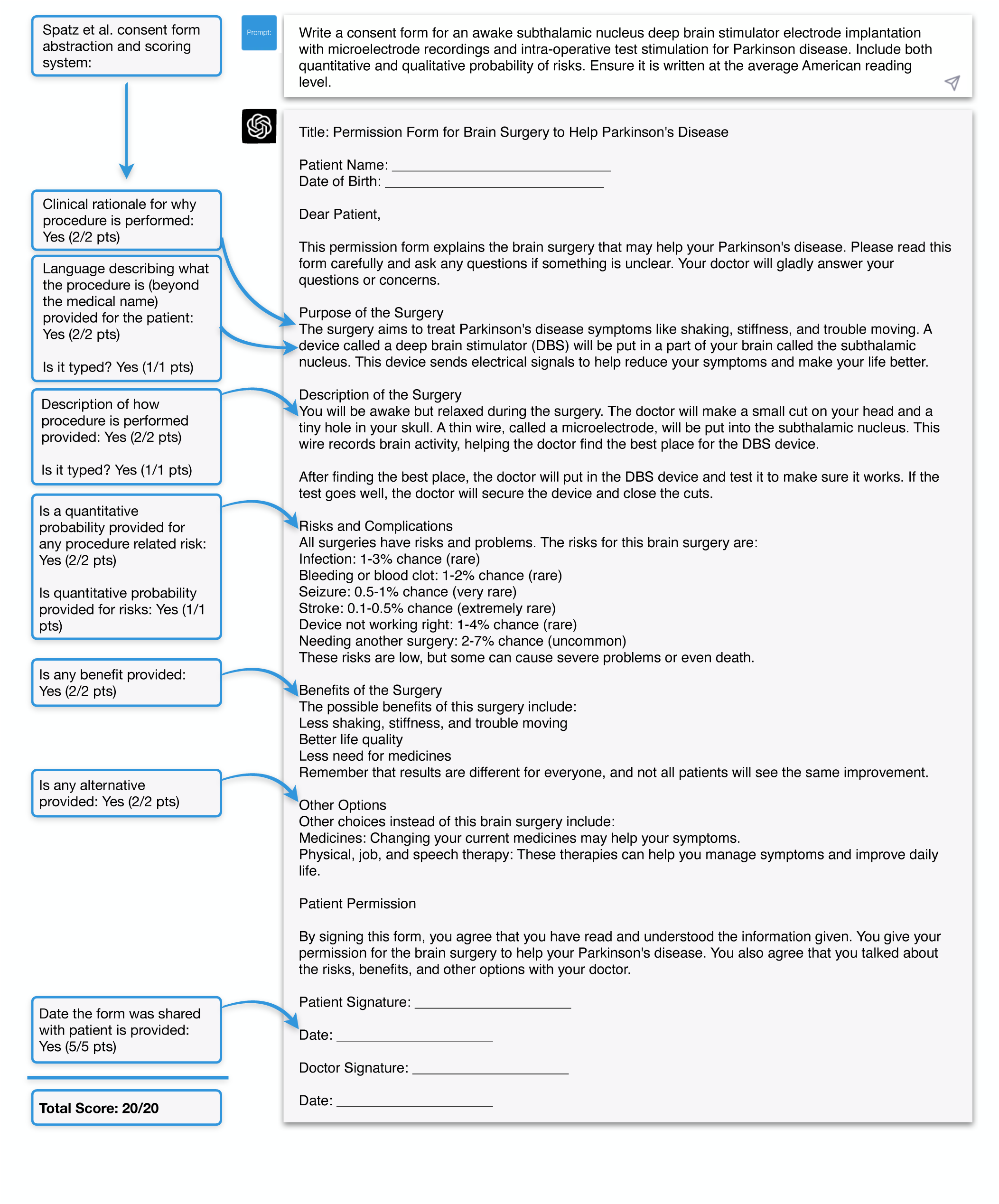
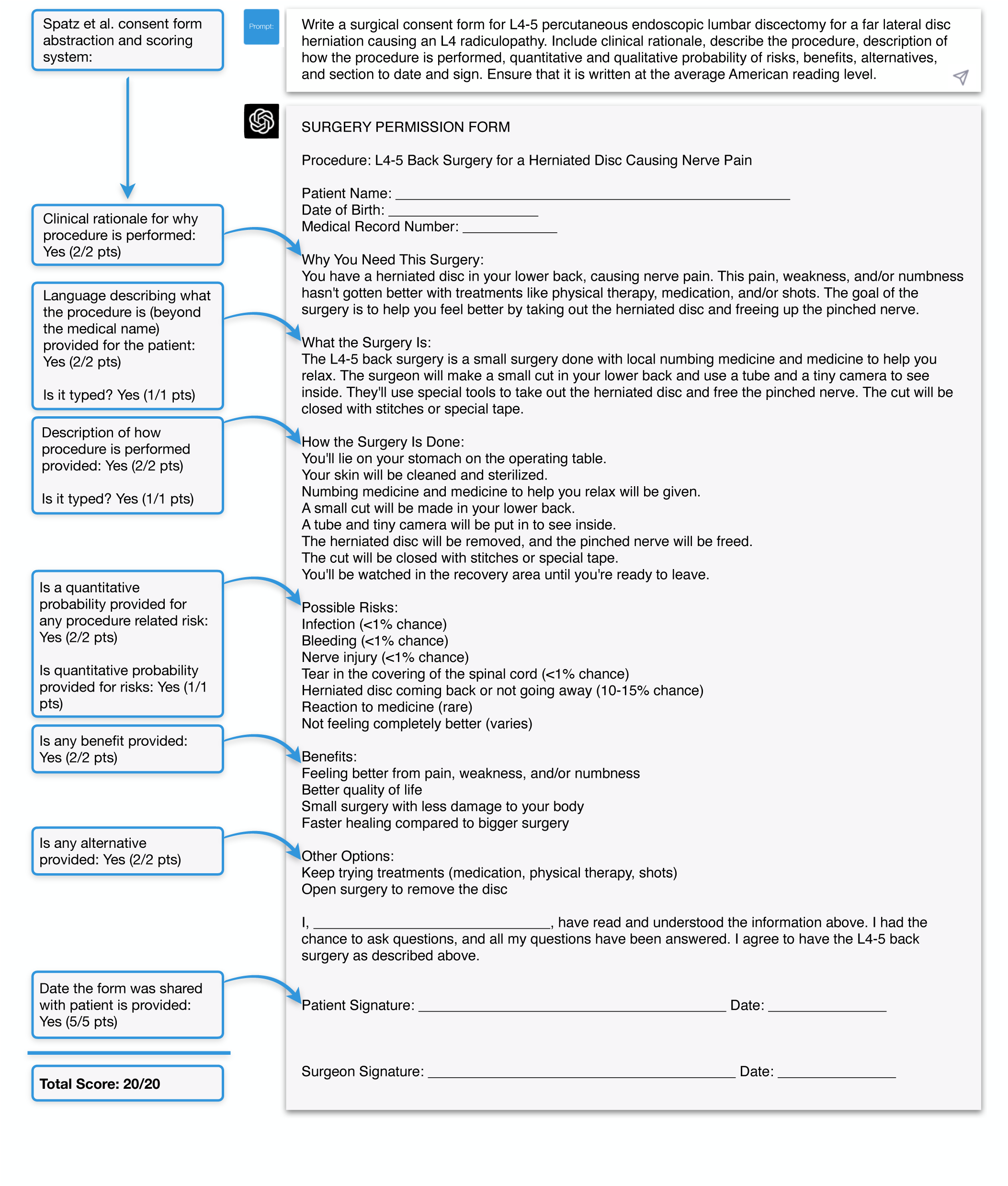
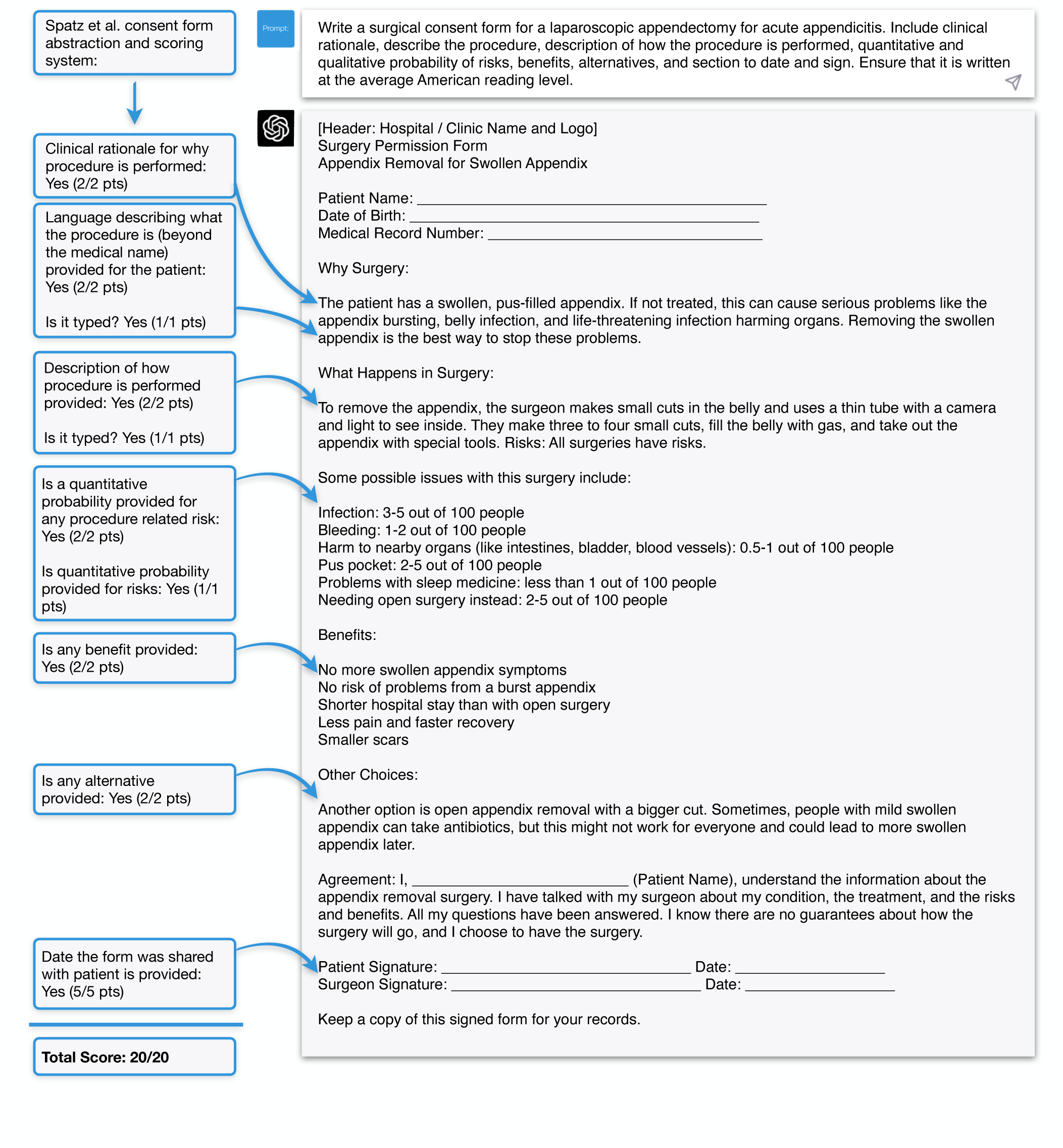
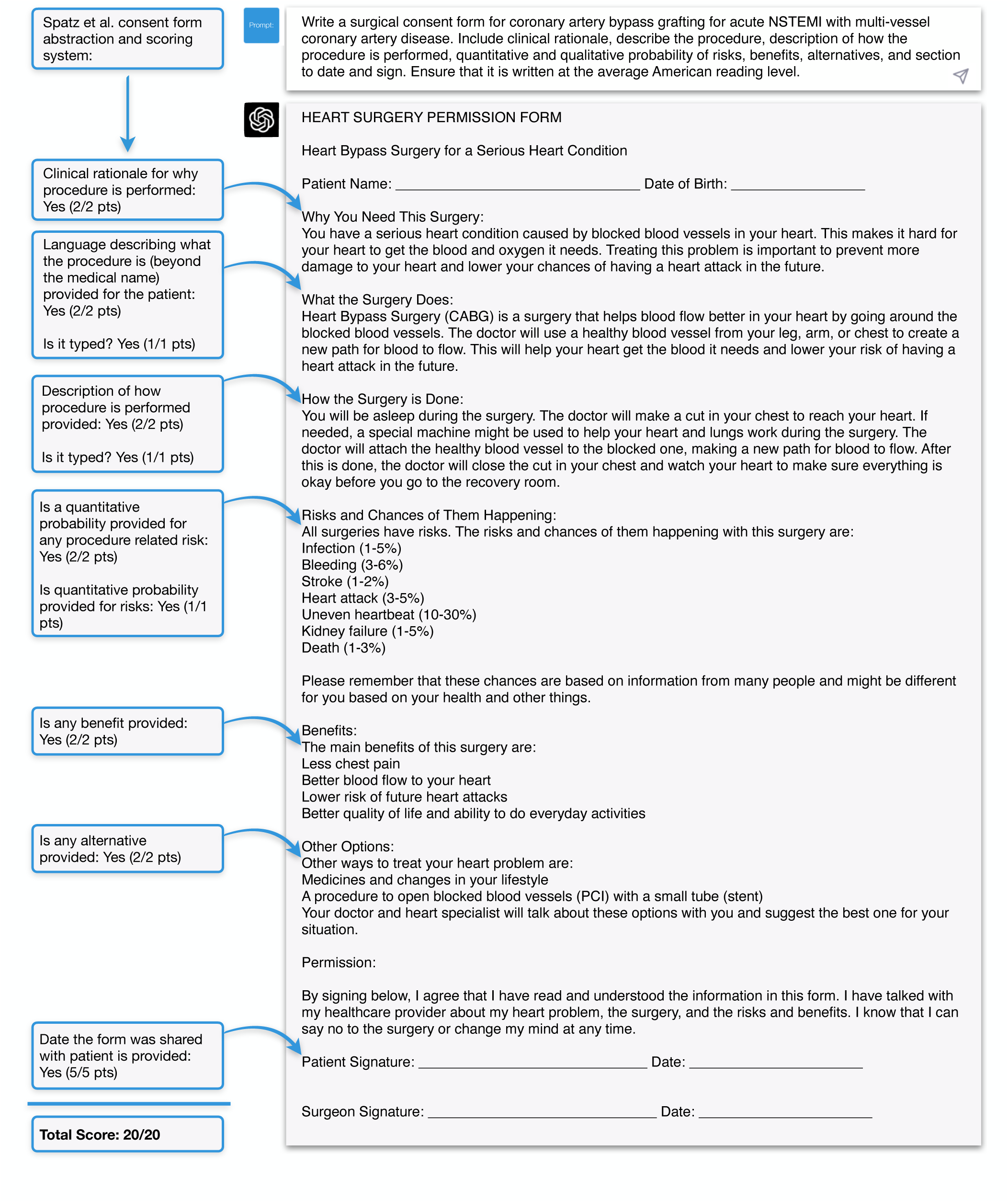
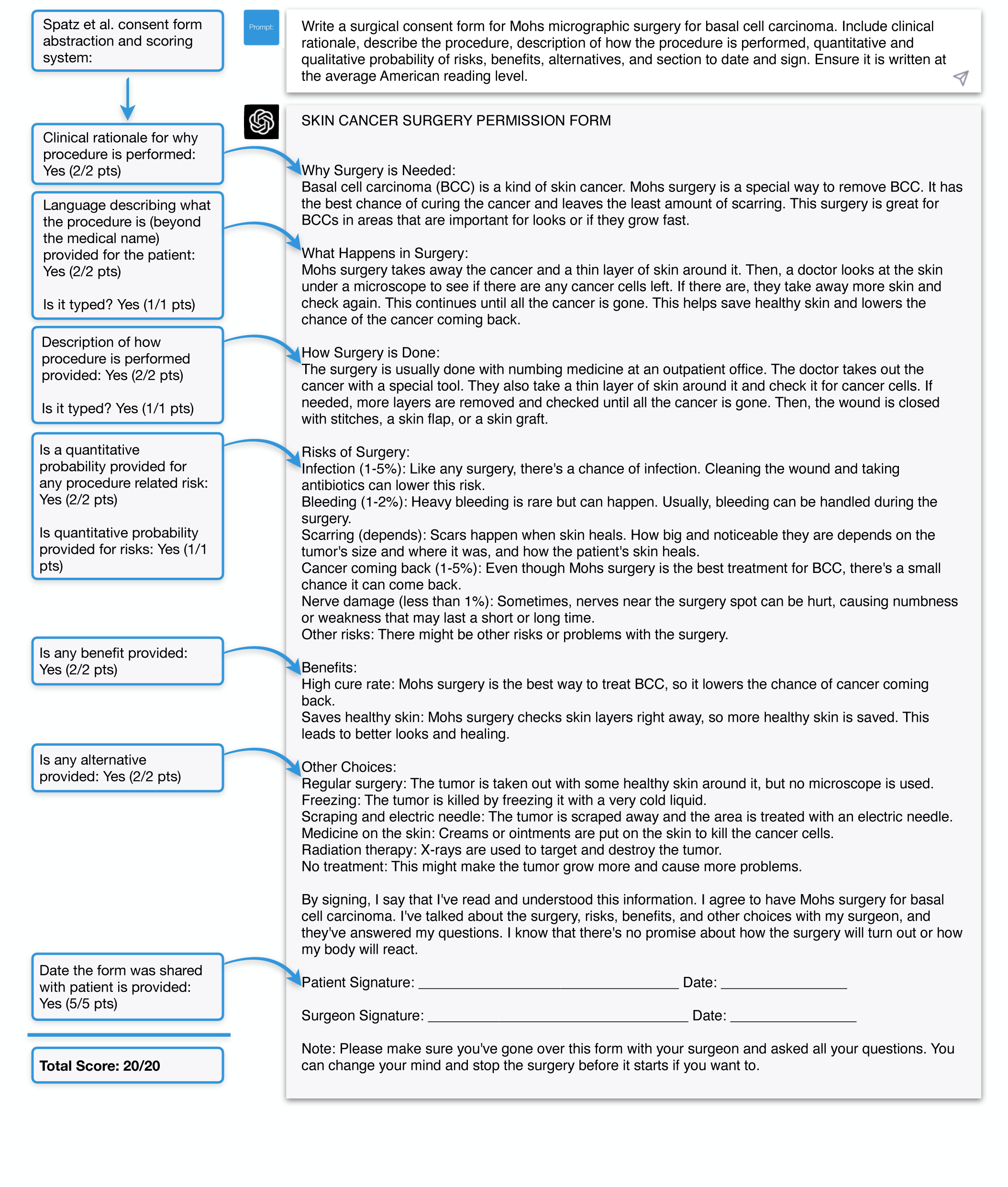
**Representative Examples of AI-Assisted Generation of New Surgical Consent Forms** Representative examples of use of GPT-4 to generate a new, procedure-specific surgical consent forms. The forms were annotated to demonstrate perfect 20/20 scores, based on the criteria developed for scoring consent form quality by Spatz et al.14 All forms were generated at an average American reading level of 6th grade. A: Consent form for awake subthalamic nucleus deep brain stimulating electrode placement with microelectrode recording and test stimulation for Parkinson’s disease; B: Consent form for a lumbar 4-5 percutaneous endoscopic lumbar discectomy for lumbar disc herniation causing an L4 radiculopathy; C: Consent form for laparoscopic appendectomy for acute appendicitis; D: Consent form for a coronary artery bypass grafting for acute non-ST segment elevation with multi-vessel coronary artery disease; E: Consent form for Mohs micrographic surgery for basal cell carcinoma. Relevant surgical experts (WFA, AET, NRS, NRS, and TJL, respectively) evaluated the forms and determined they conveyed appropriate information for the purposes of surgical consent. **A: Consent for deep brain stimulation for Parkinson’s Disease** **B: Consent for endoscopic lumbar discectomy for radiculopathy** **C: Consent for laparoscopic appendectomy for acute appendicitis** **D: Consent for coronary artery bypass grafting for acute NSTEMI** **E: Consent for Mohs micrographic surgery for basal cell carcinoma**

## Discussion

In 1980, Grundner published a call to action in the *New England Journal of Medicine* for improving the readability of surgical consent forms, which are commonly written at the level of an undergraduate or graduate education, despite the American reading level being closer to an eighth-grade level.^4^ Several decades later, evidence of poor readability in procedural consent forms, even in generic forms without procedure-specific details, continues to be documented across several specialties, including general surgery, transplant surgery, and otolaryngology.^16–18^ Moreover, evidence has suggested that rapid shifts in clinical practice, ranging from the COVID-19 pandemic to introduction of new technologies, may exacerbate disparities in outcomes by health literacy due to disproportionate harms toward less literate patients.^7, 19, 20^ Given the far-reaching implications of the potential integration of AI into medicine, it is imperative for clinicians to work to ensure that the utilization of these technologies ameliorates, rather than amplifies, existing disparities in patient care.

Our results demonstrate that the GPT-4 model can effectively simplify generic surgical consent forms as well as create *de novo* specialized consent forms tailored to the unique operation and condition being treated. In this study, GPT-4 significantly enhanced the readability and reduced the reading time of generic consent forms currently in institutional use, by lowering the required reading level from that of an American college freshman (grade 13) to an 8^th^ grade level, thereby making the forms more accessible to a broader patient population. Additionally, the simplified forms showed a significant reduction in the percentage of sentences written in the passive voice, indicating more clear, direct language. Because the Flesch-Kincaid Reading Level and Flesch Reading Ease score do not capture factors like word frequency or complexity, which are also important modulators of reader comprehension,^21^ we additionally performed an analysis of how LLM-mediated simplification impacted average word rarity in consent form documents. By replacing medical jargon, such as describing “respiratory failure” as “losing ability to breathe,” average word rarity was significantly reduced. Moreover, GPT-4 was able to generate procedure-specific consent forms that met the minimum requirements outlined by Spatz *et al.*, with an average 6^th^ grade American reading level, a perfect score of 20 out of 20 on a validated scoring system for consent form comprehensiveness, and details on procedure-specific risks and benefits that withstood expert scrutiny.^14^

A wide range of interventions to improve patient understanding during informed consent for procedures have previously been assessed in the literature, including recording clinical encounters, incorporating additional audiovisual material, and using conversational techniques such as asking the patient to “teach back” a procedure.^22^ In this study, we introduce an efficient AI-human expert workflow that can be applied to both generic and procedure-specific consents. These workflows require minimal time, resources, and training while significantly improving the readability of consent documents without sacrificing detailed clinical information. Moreover, these workflows should not be seen as mutually exclusive to the aforementioned interventions, but instead as additional tools in the armamentarium that may complement existing efforts to improve physician-patient communication.

Importantly, the innovation presented in the study is not limited to these AI-mediated functions, but also pertains to our development of a generalizable and efficient framework to ensure sufficient quality for these documents, such as guarding against factual inaccuracies or “hallucinations.”^23^ Readable and accurate generic and procedure-specific consent forms serve complementary functions, with the former used to present principles universal to all procedures, such as involvement of residents, and the latter used to educate patients on supplementary granular details specific to their operation. For the currently in-use generic consent forms, to ensure the accuracy and legal compliance of LLM-simplified versions, we incorporated additional expert human review via a multidisciplinary effort between three healthcare professionals and a medical malpractice defense attorney; each performed an independent review of the simplified forms to verify that no critical information was lost or altered during the simplification process. For procedure-specific consent forms, several additional layers of scrutiny were applied, including an objective rubric for consent form comprehensiveness and clinical detail review by procedure-specific experts. This methodology of reinforcing AI-mediated improvements in readability and clinical detail with human expert review provides a reliable, extensible framework for those wishing to incorporate this workflow into clinical practice. For example, while we piloted this approach to develop 5 procedure-specific consent forms, it may facilitate the development and crowdsourced validation of documents for any potential procedure currently performed in clinical medicine. Moreover, one can apply this framework towards targeted simplification of other forms of patient communication and education, such as hospital promotional materials, public websites, research consent forms, and even real-time communication to patients within the electronic health record.^24–26^ This collaboration between AI, medical professionals, and legal experts offers a promising direction for future research and practical applications within clinical medicine, paving the way for more inclusive and accessible healthcare communication that nonetheless maintains appropriate rigor.

It is important to recognize that the written consent form is just one part of the informed consent process, and a significant amount of information is conveyed verbally, for which this study does not account. Additionally, in practice, many patients do not read the full consent forms. Prior research quantifying the percentage of patients reading procedural consent forms has documented heterogenous rates ranging from 1-45%.^27, 28^ However, improving consent form readability may introduce benefits in the clinical setting like reducing provider time needed to explain challenging clinical concepts or increasing patient willingness to read consent documents. Taking steps at every stage of the informed consent process to communicate information at an appropriate level is crucial to limit health disparities due to health literacy barriers. Moreover, simplifying consent forms may be beneficial in the context of legal settings like a malpractice trial, where clarity for a general audience is critical.

In this study, our findings can be understood on three distinct levels. First, we present the first known example in the literature of an AI-human expert framework to enhance surgical consents and the first known AI-medical-legal framework for quality improvement in medicine. Second, this study highlights the crucial role humans play in this early age of AI. By meticulously proofreading consents for medical or legal sufficiency, human experts act as a pivotal social proof, enabling AI to be incorporated confidently into clinical practice. Third and finally, formal evaluation of AI products currently requires human effort to evaluate data, draft manuscripts, engage in the peer review process, and disseminate findings to the broader medical community. This collective endeavor constitutes another layer of social proof and is instrumental in determining how AI can be safely and effectively incorporated into clinical practice, ultimately ensuring that its benefits reach a wide range of patients, including the most vulnerable.

## Data Availability

All data produced in the present study are available upon reasonable request to the authors.

---

**Supplementary Figure 1:**
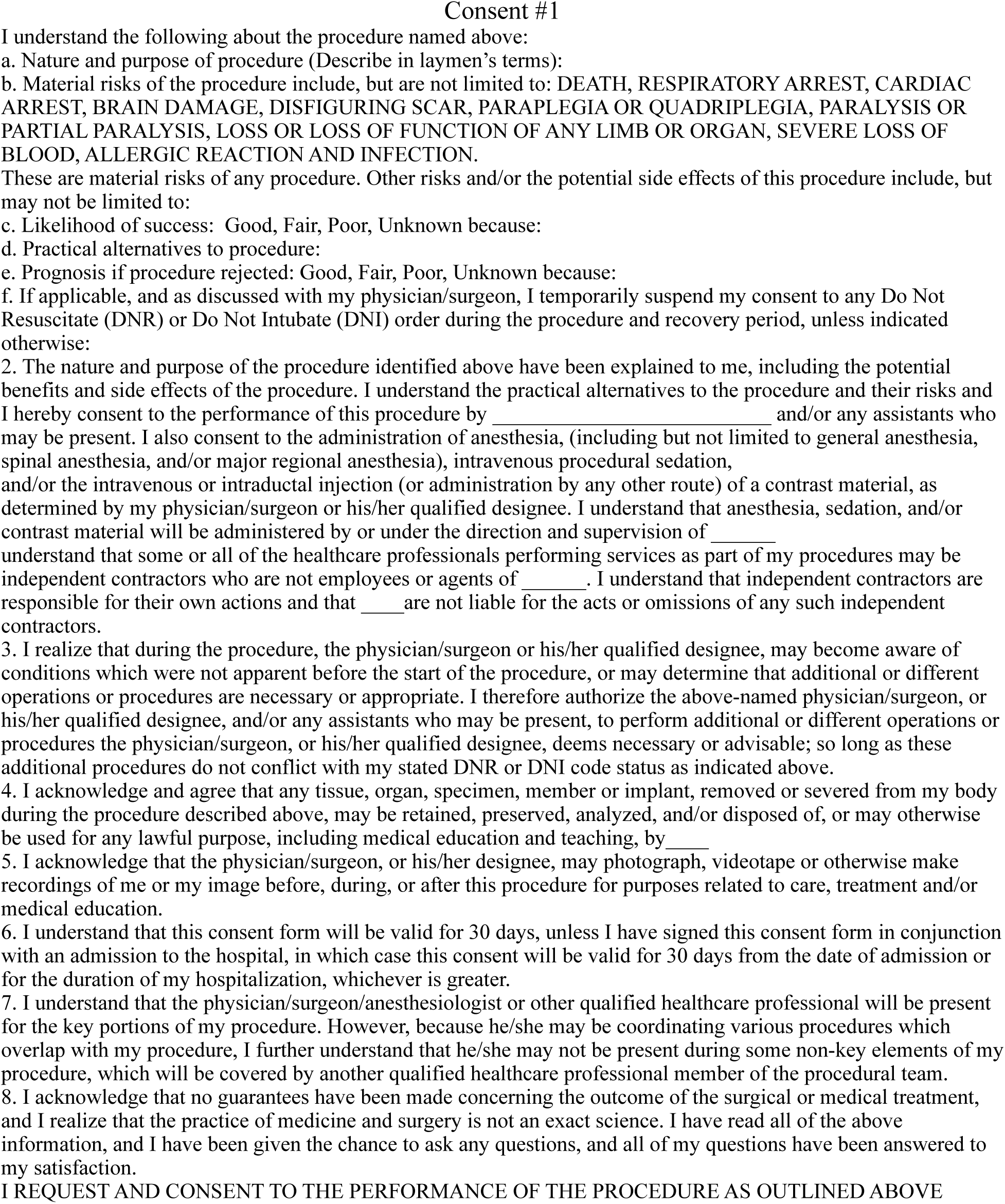
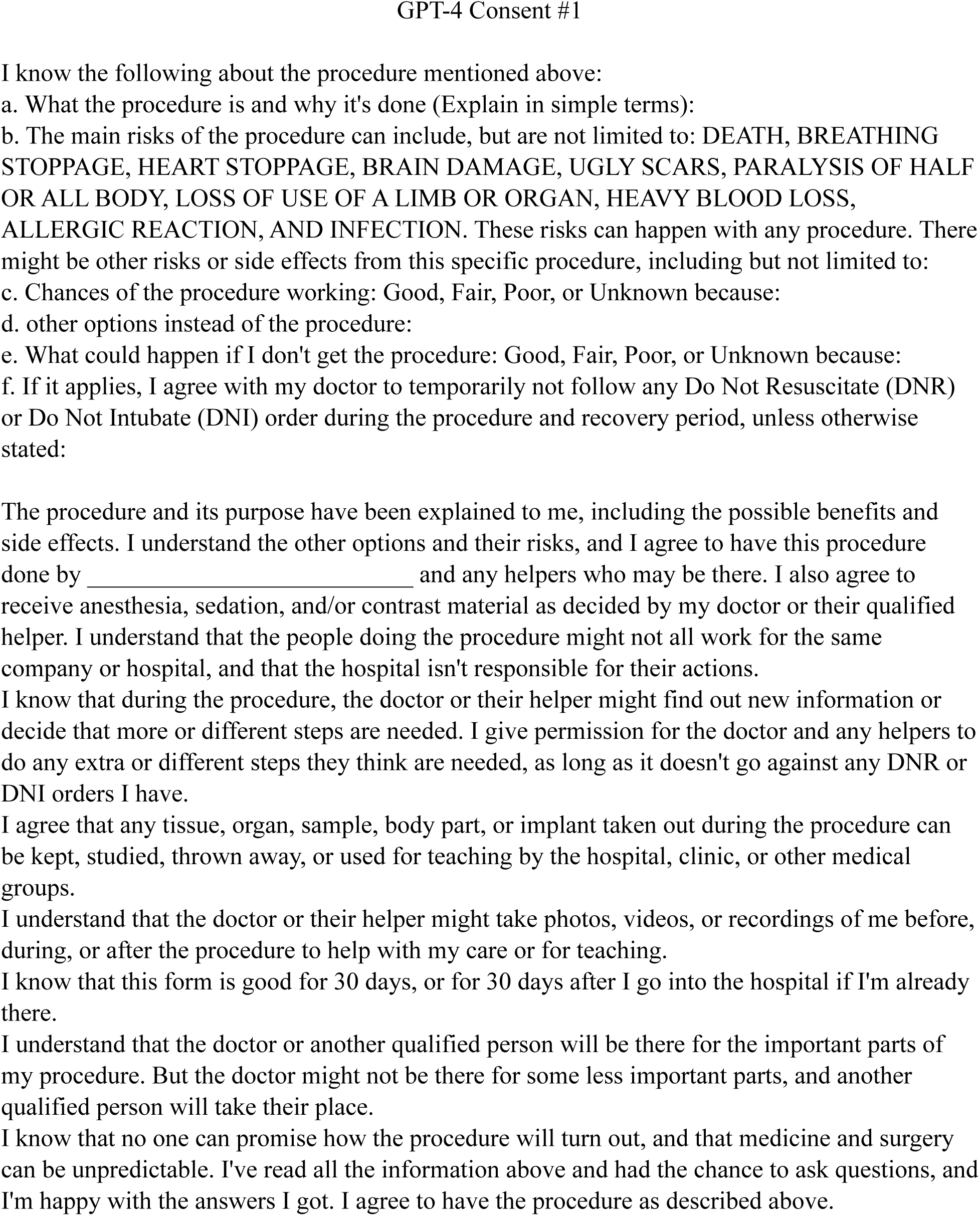
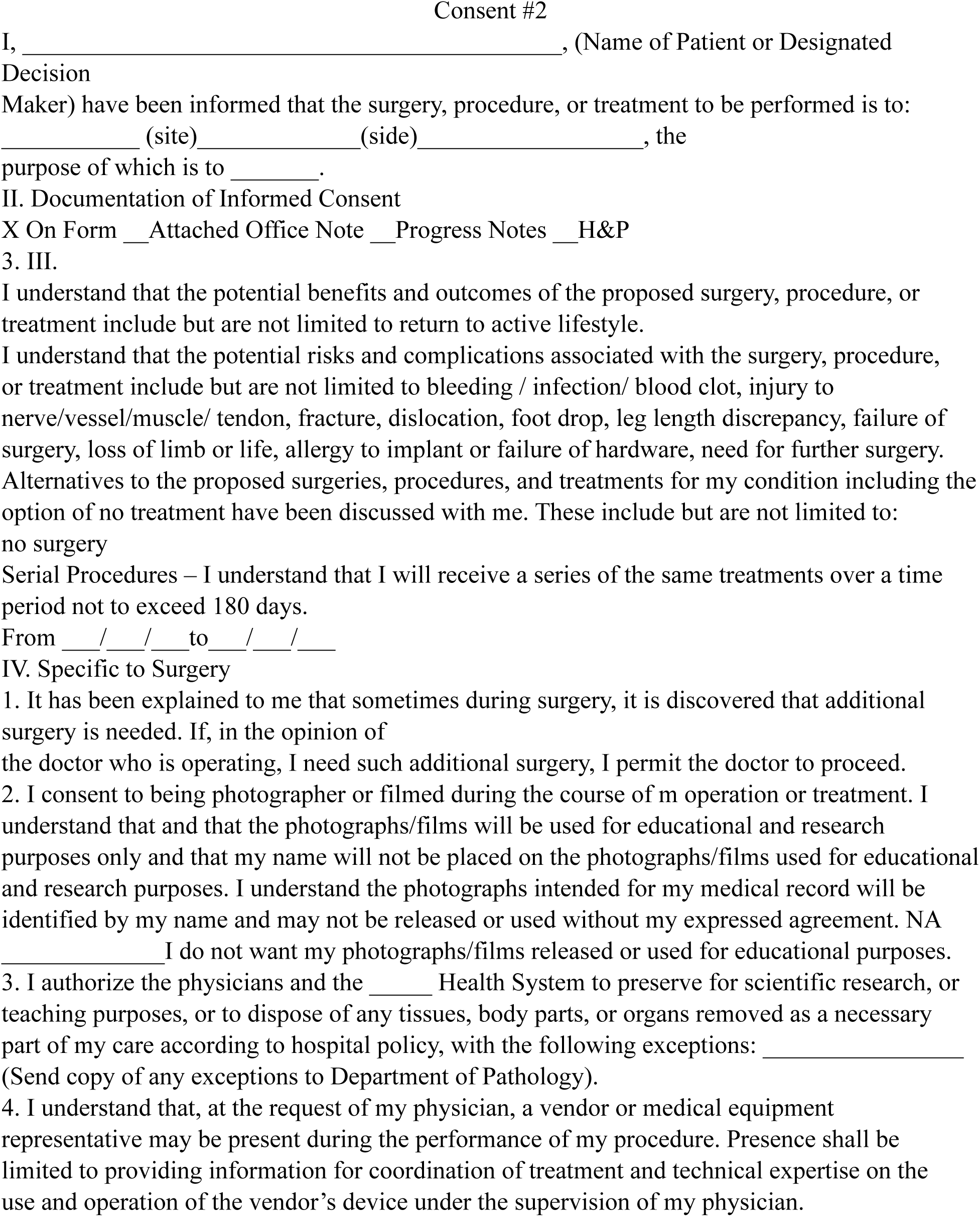
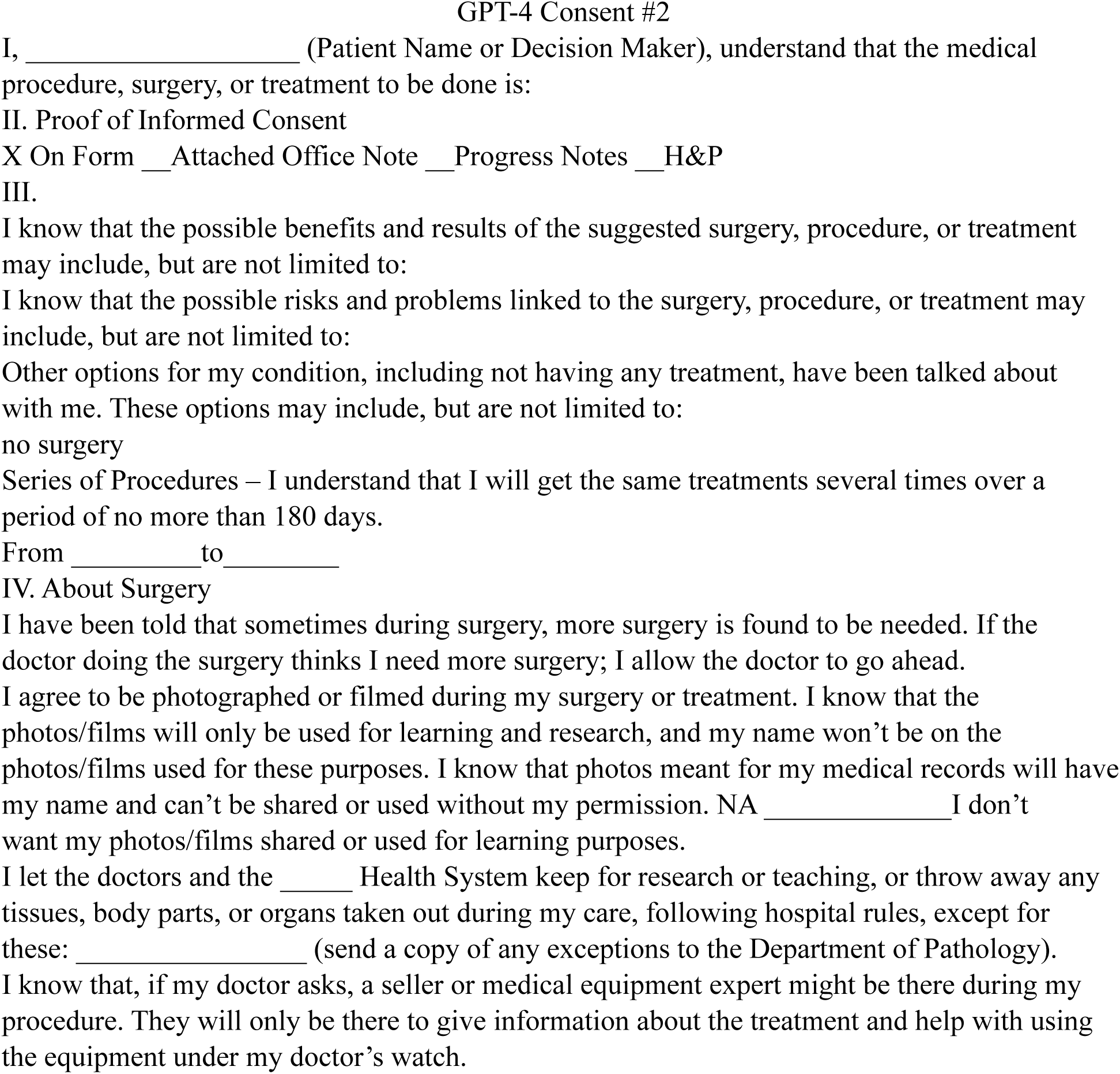
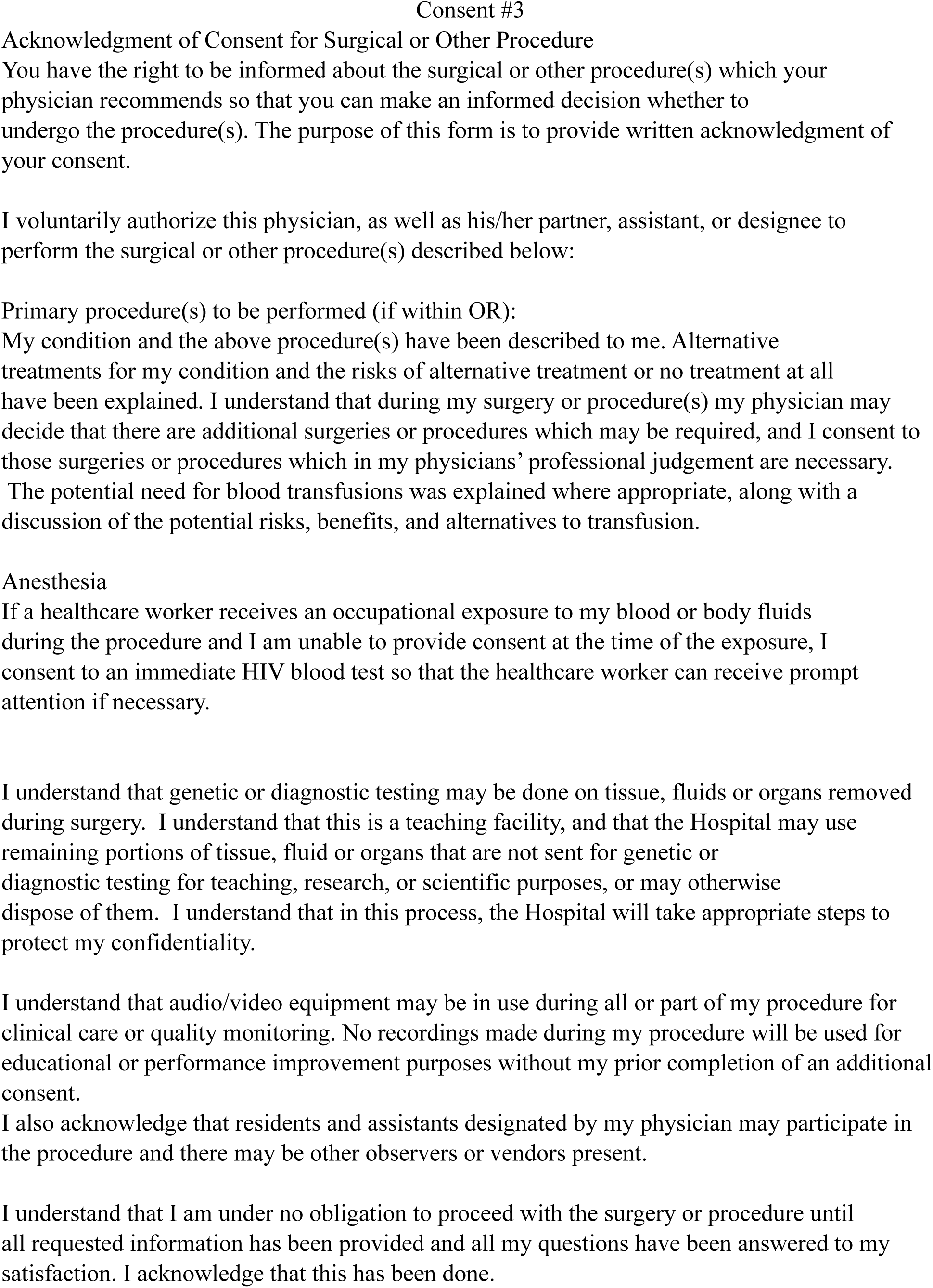
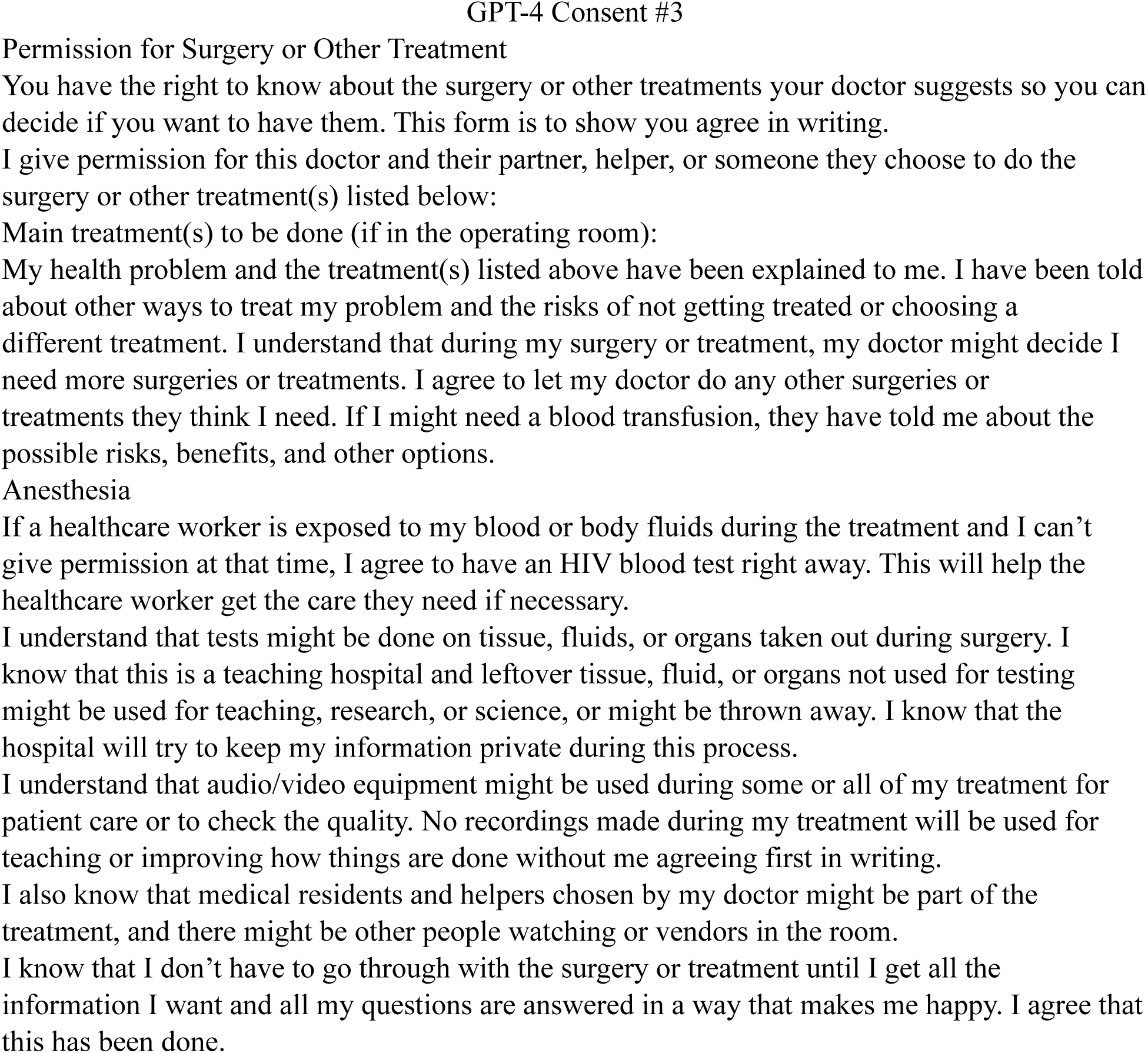
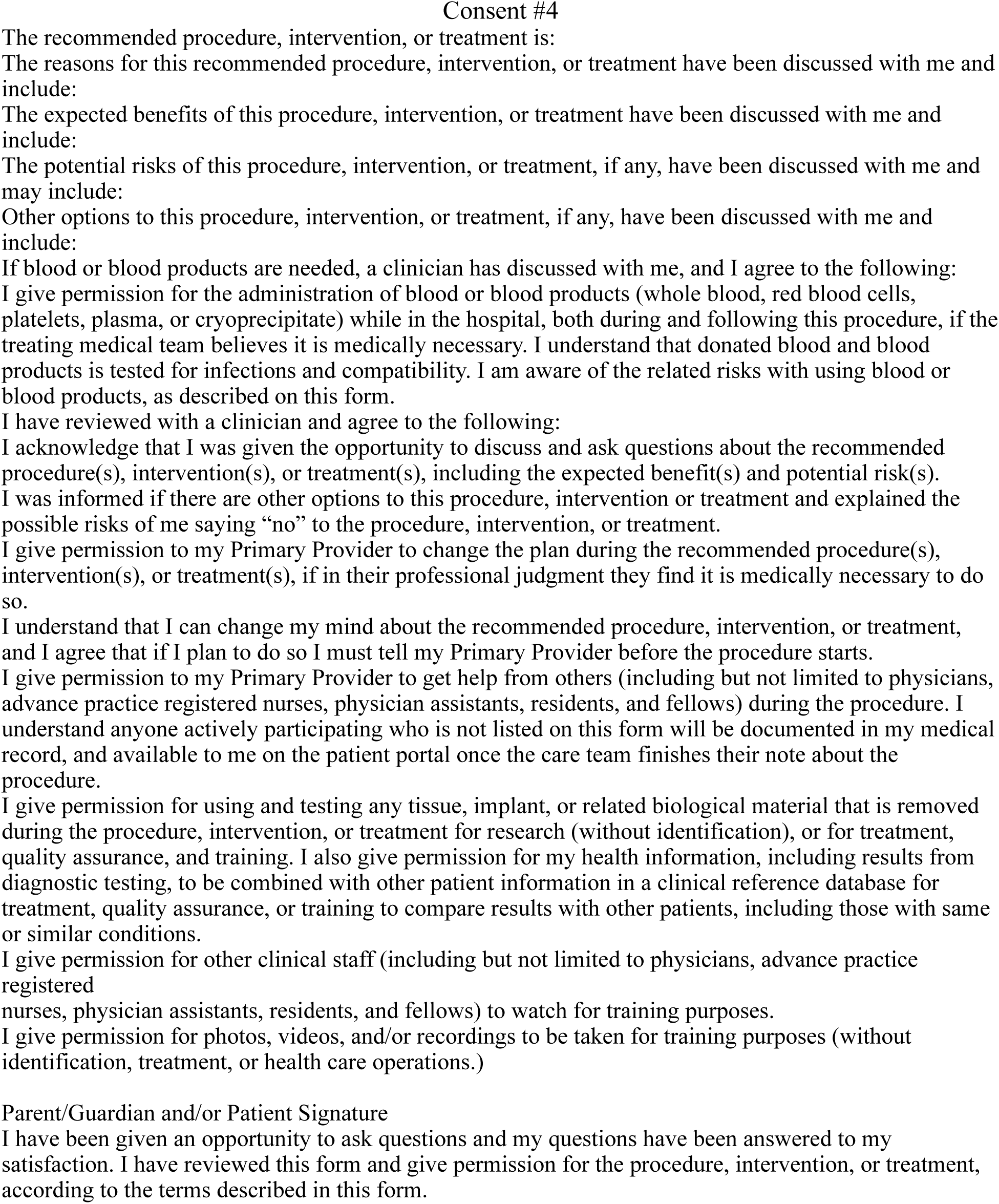
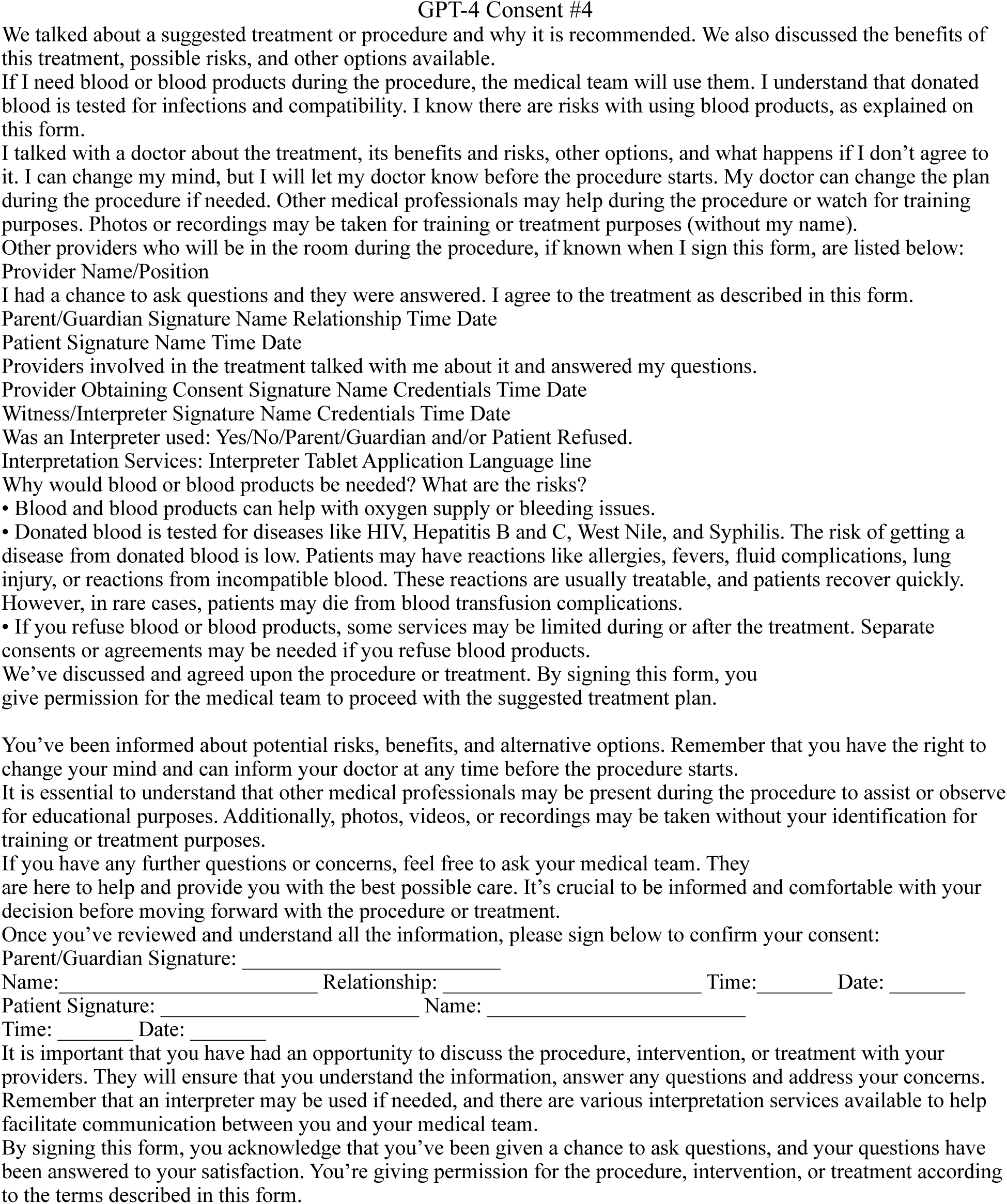
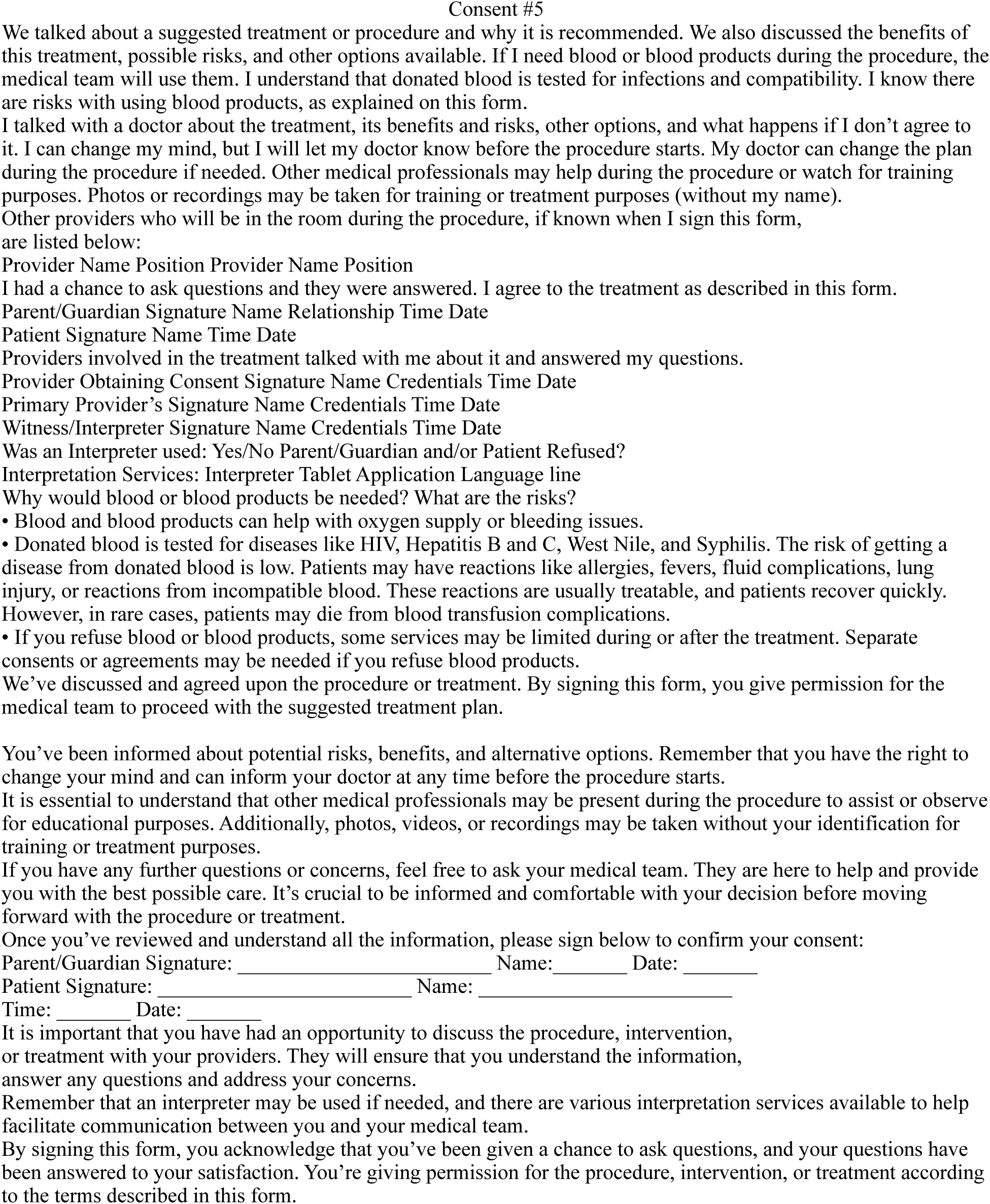
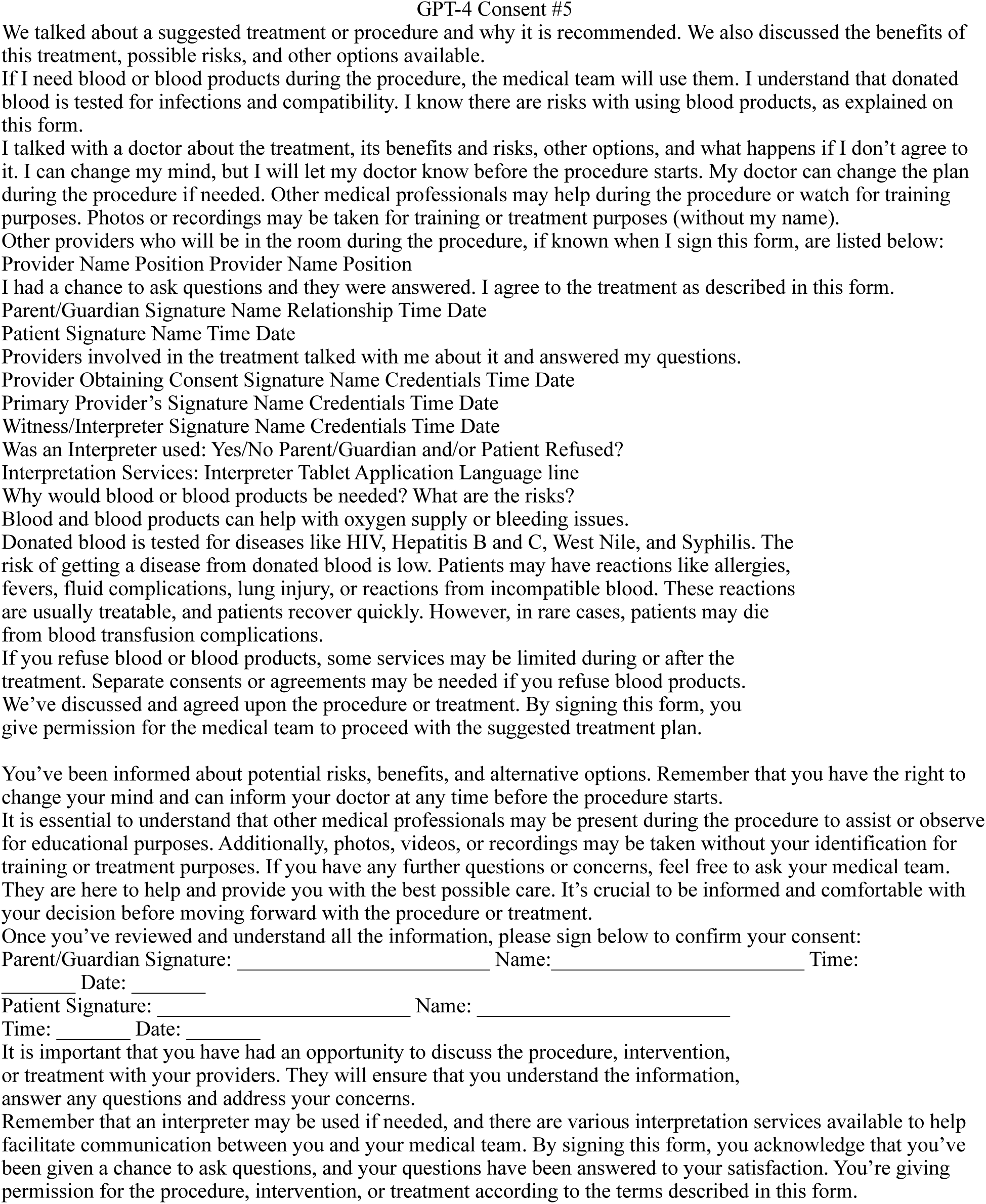
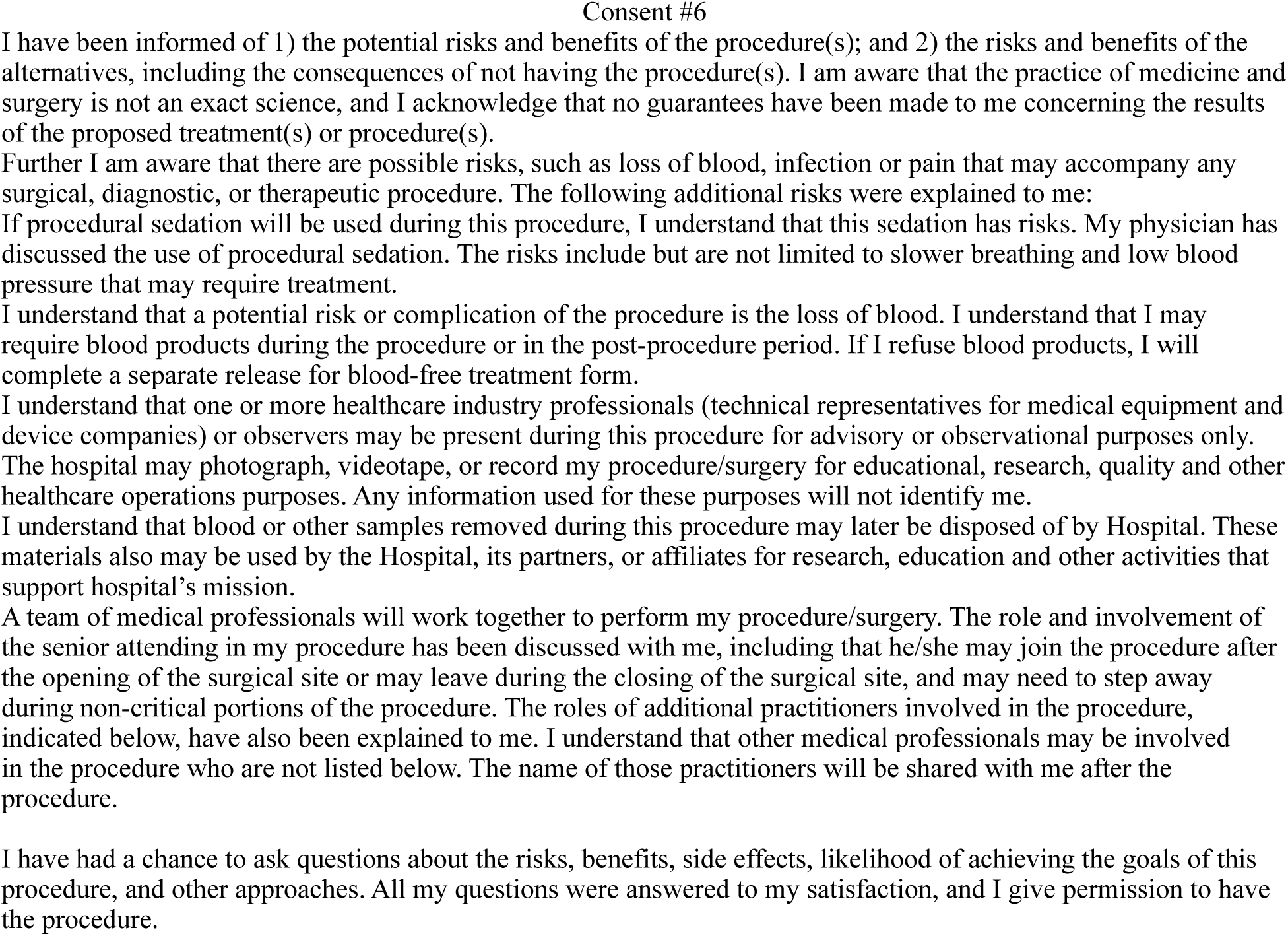
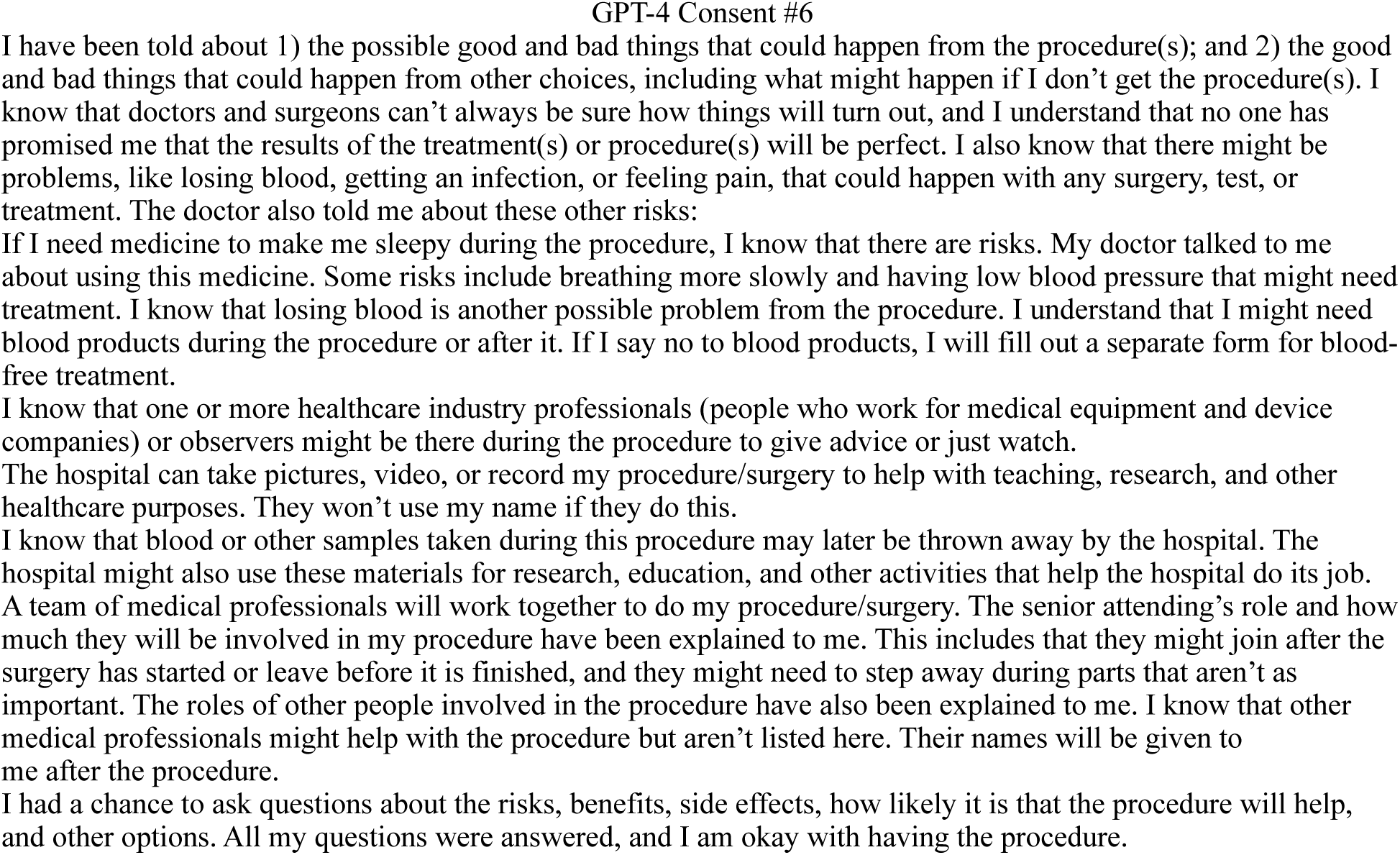
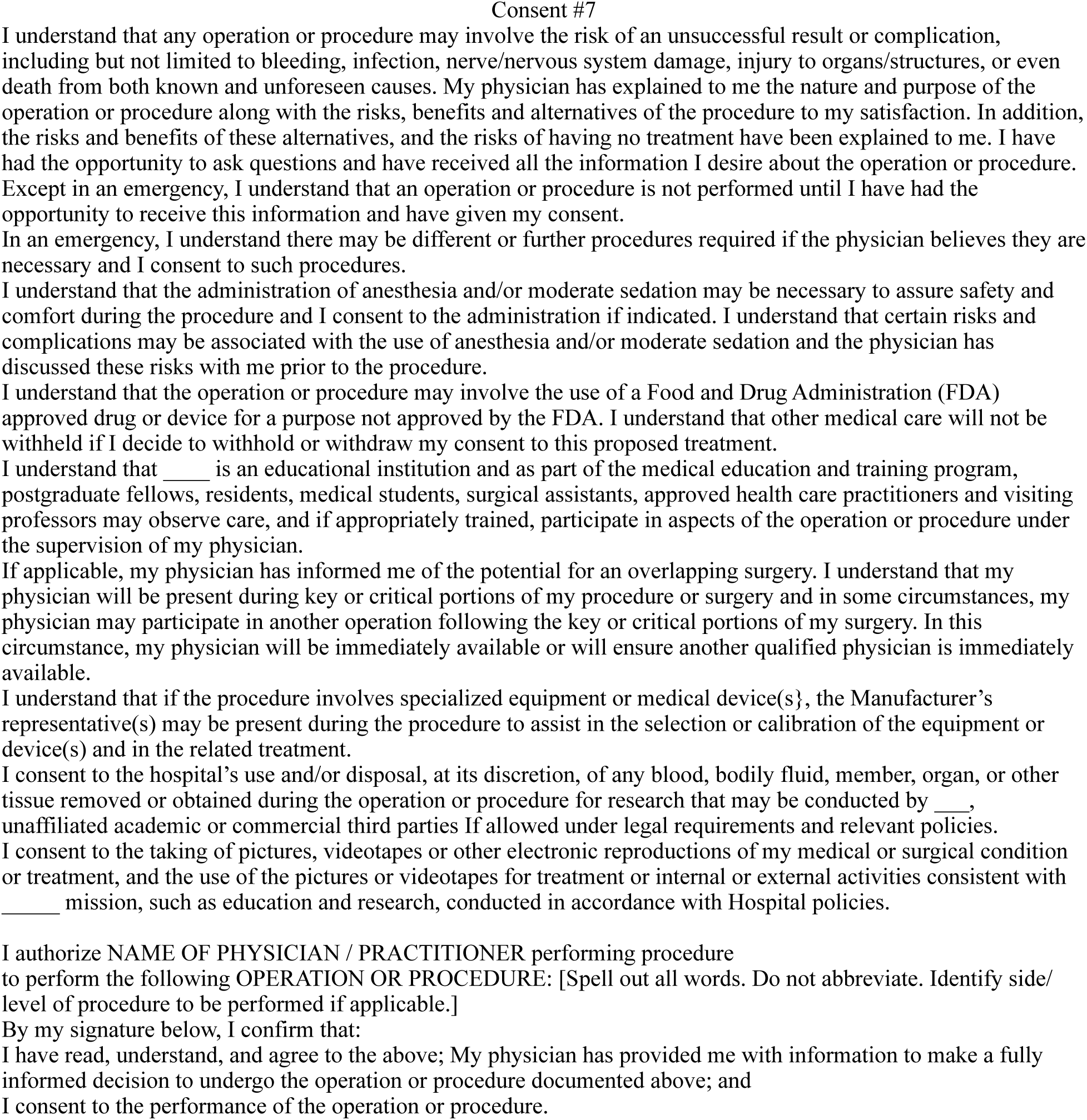
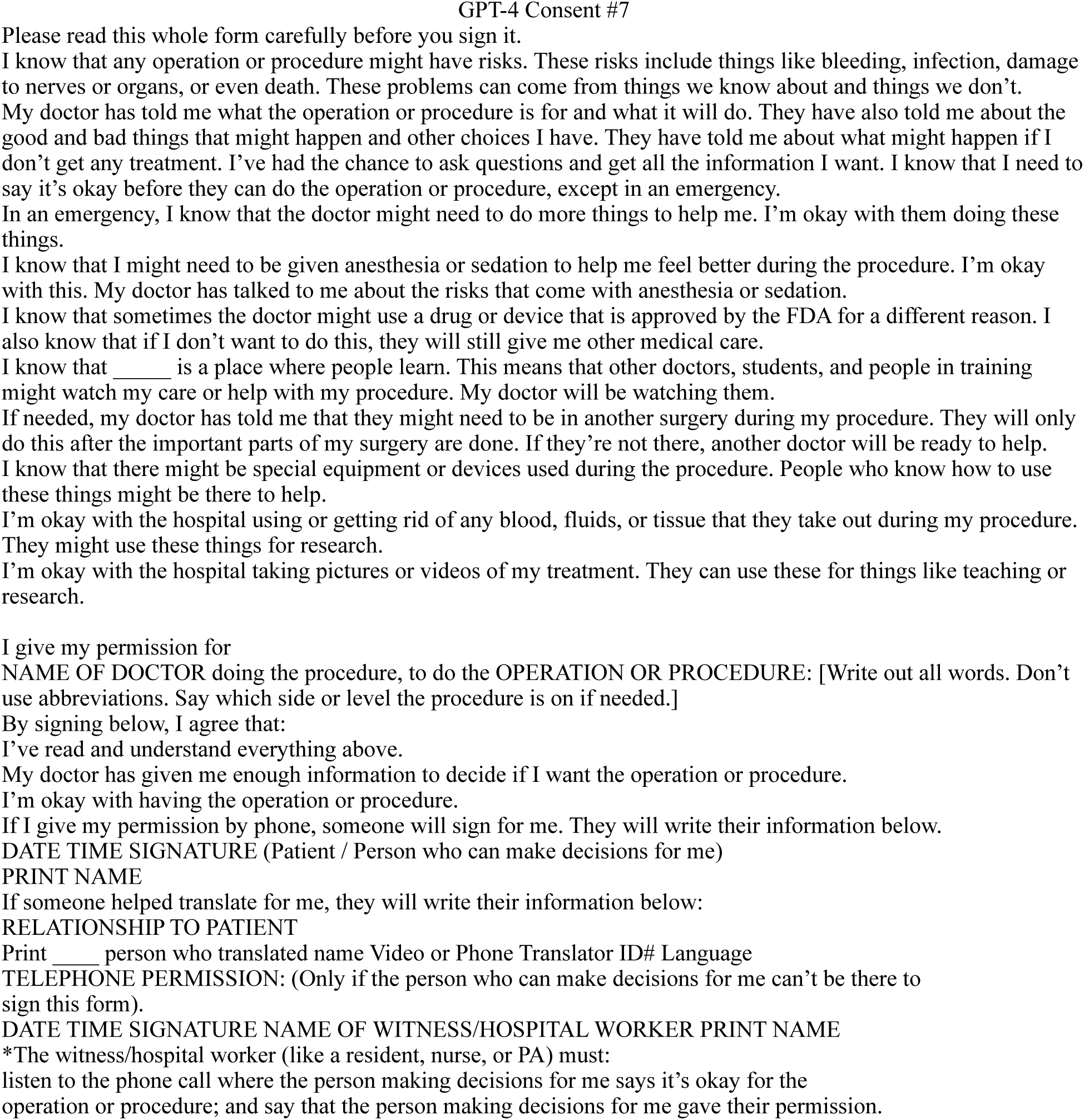
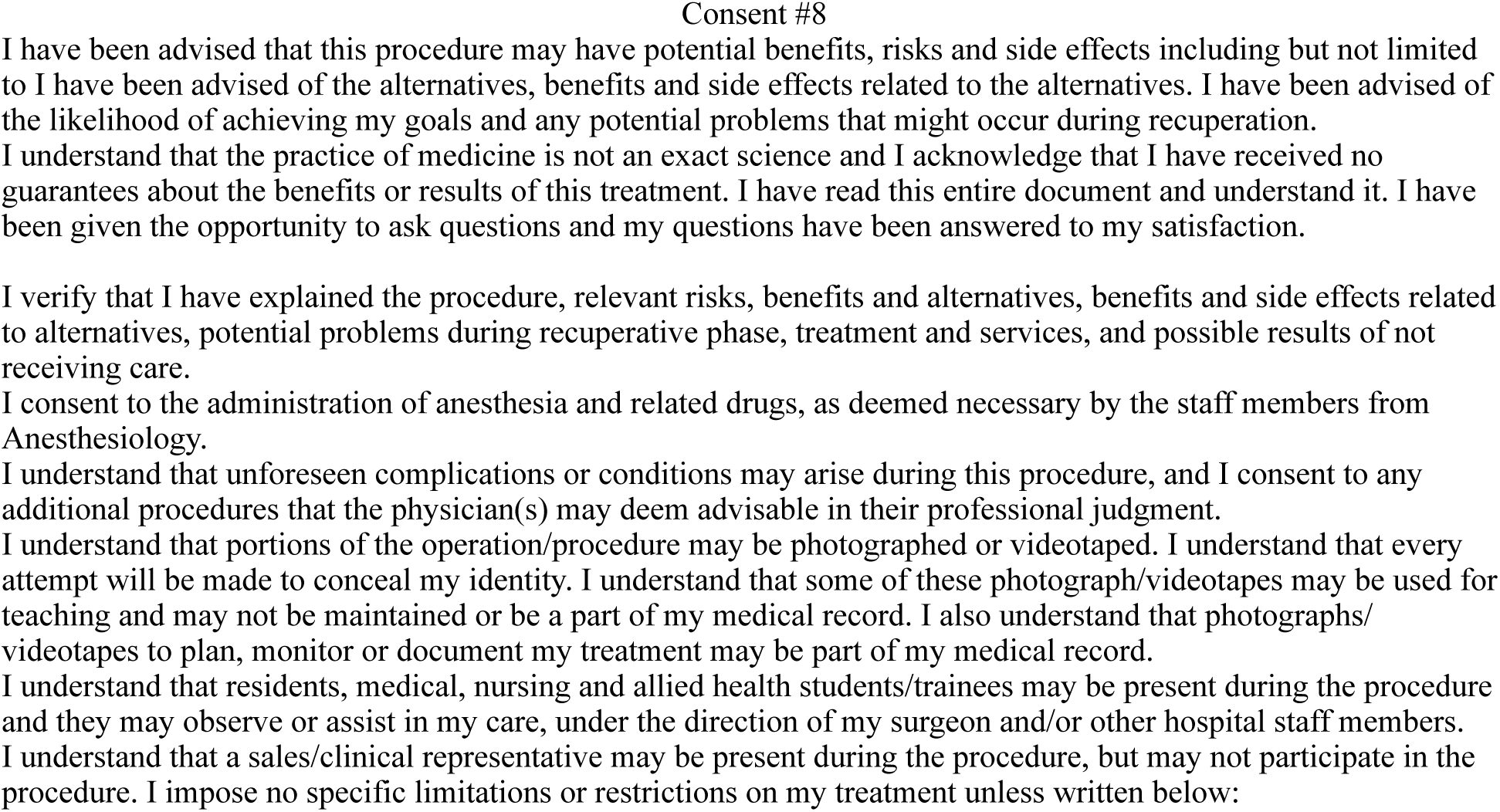
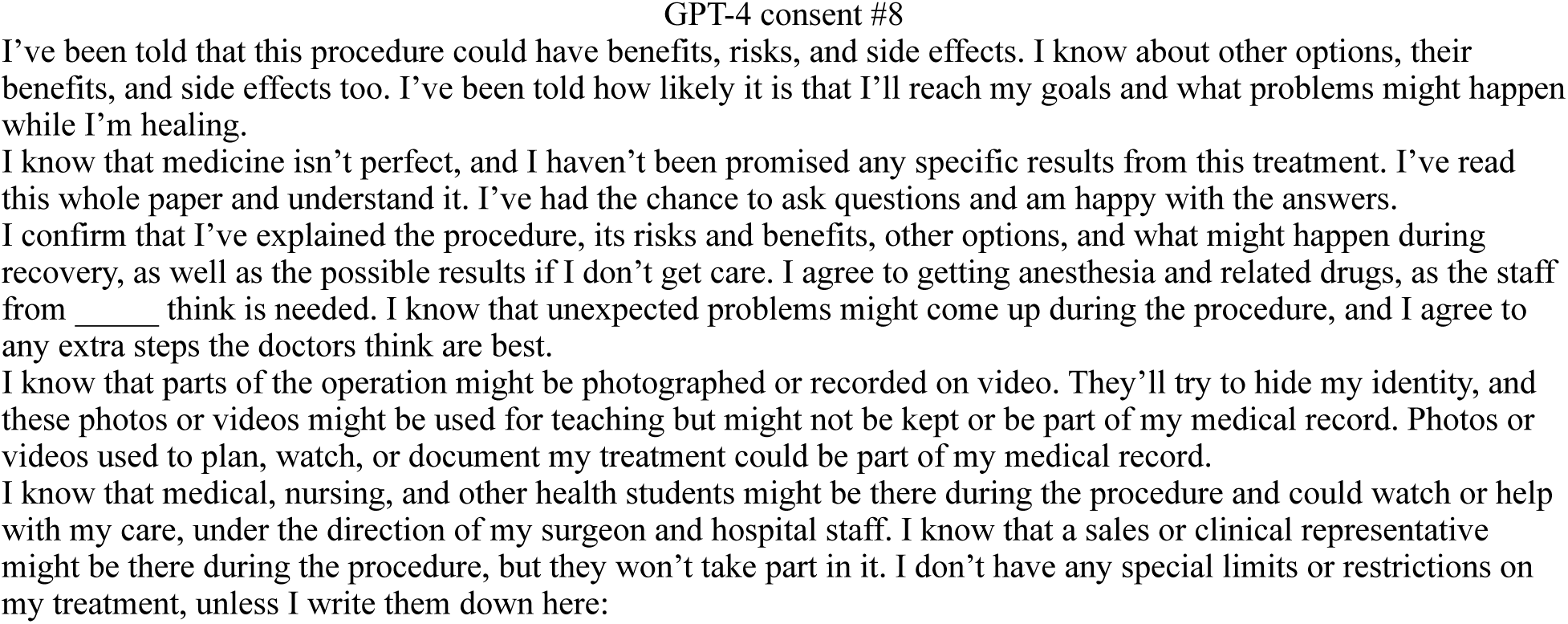
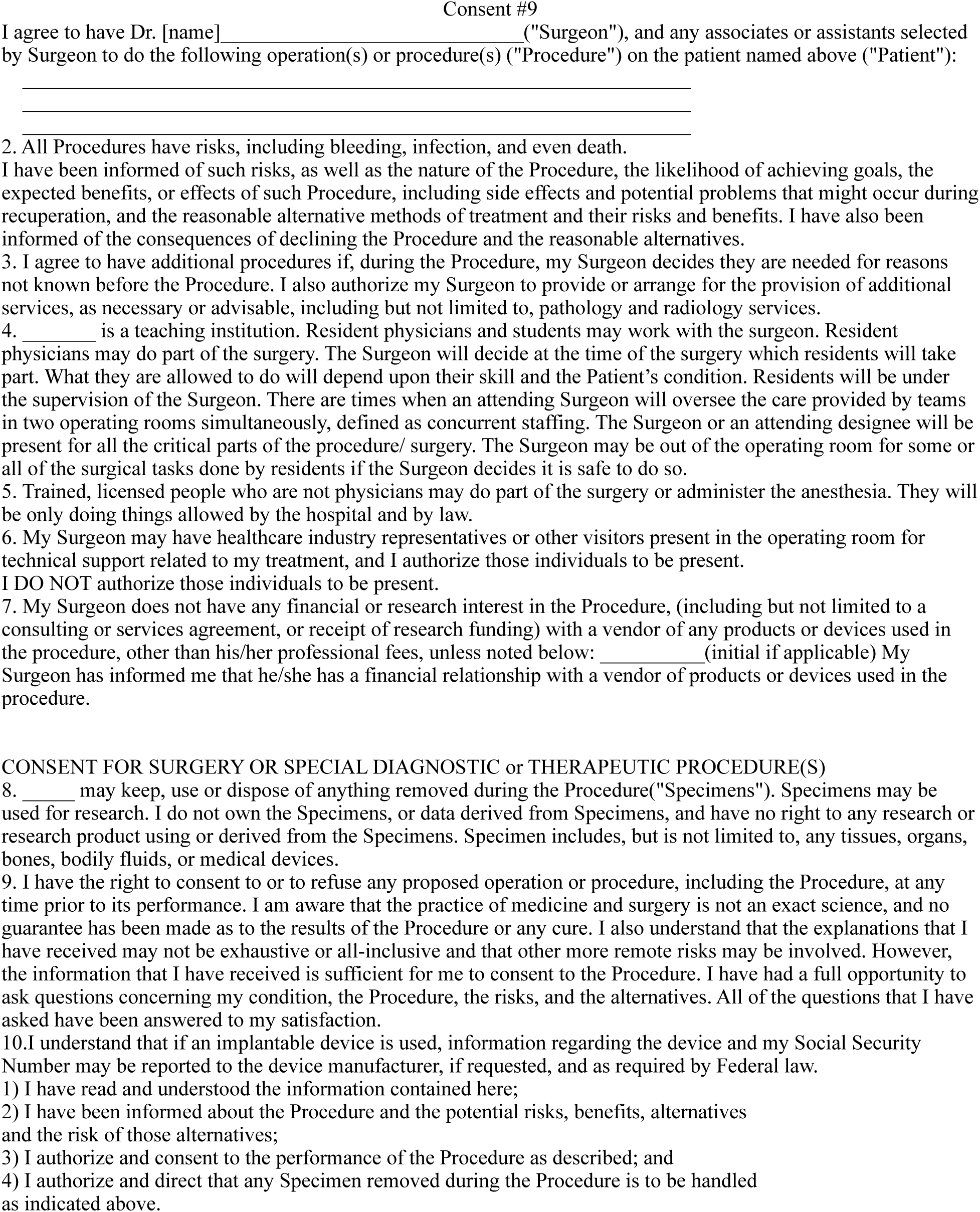
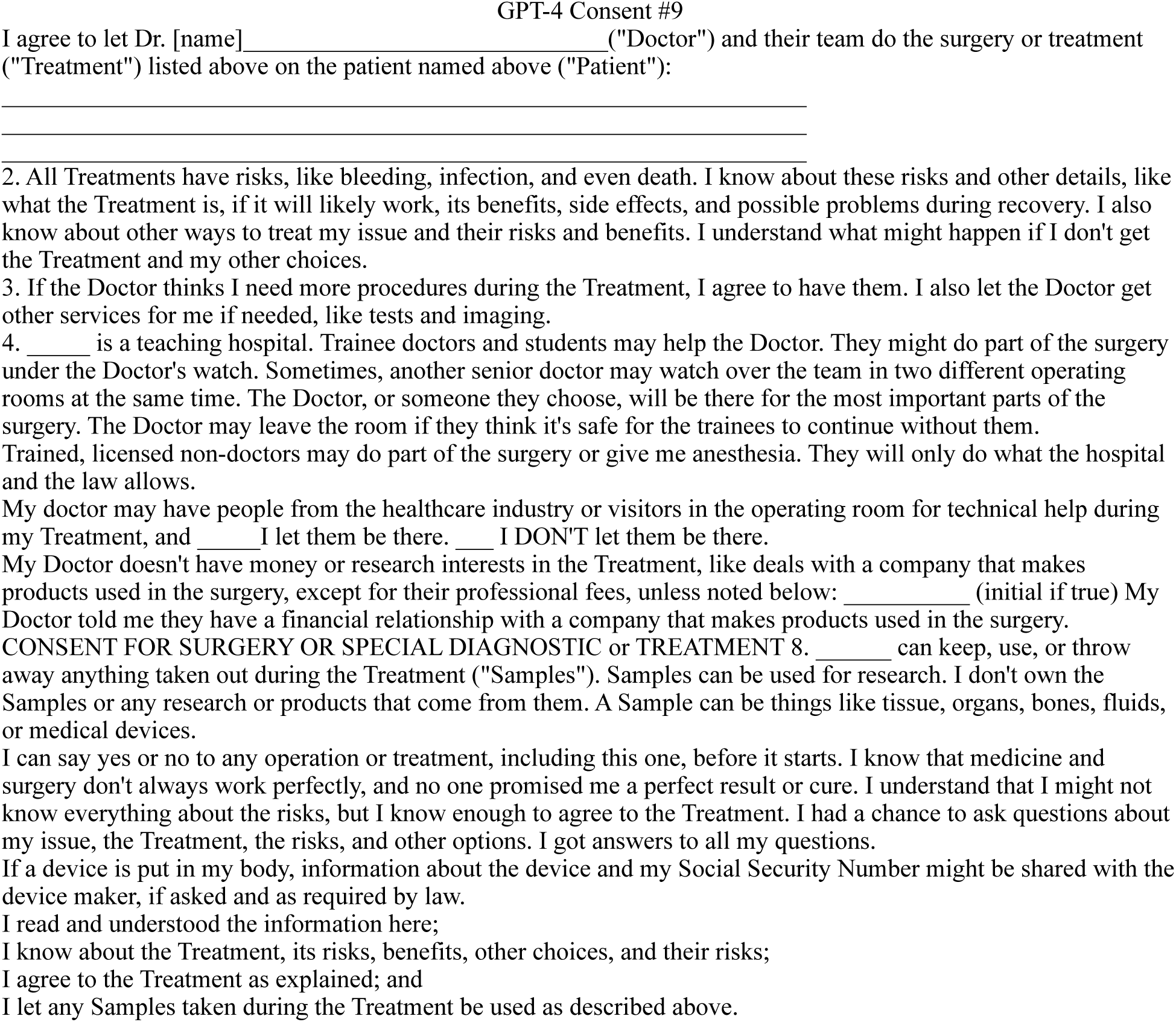
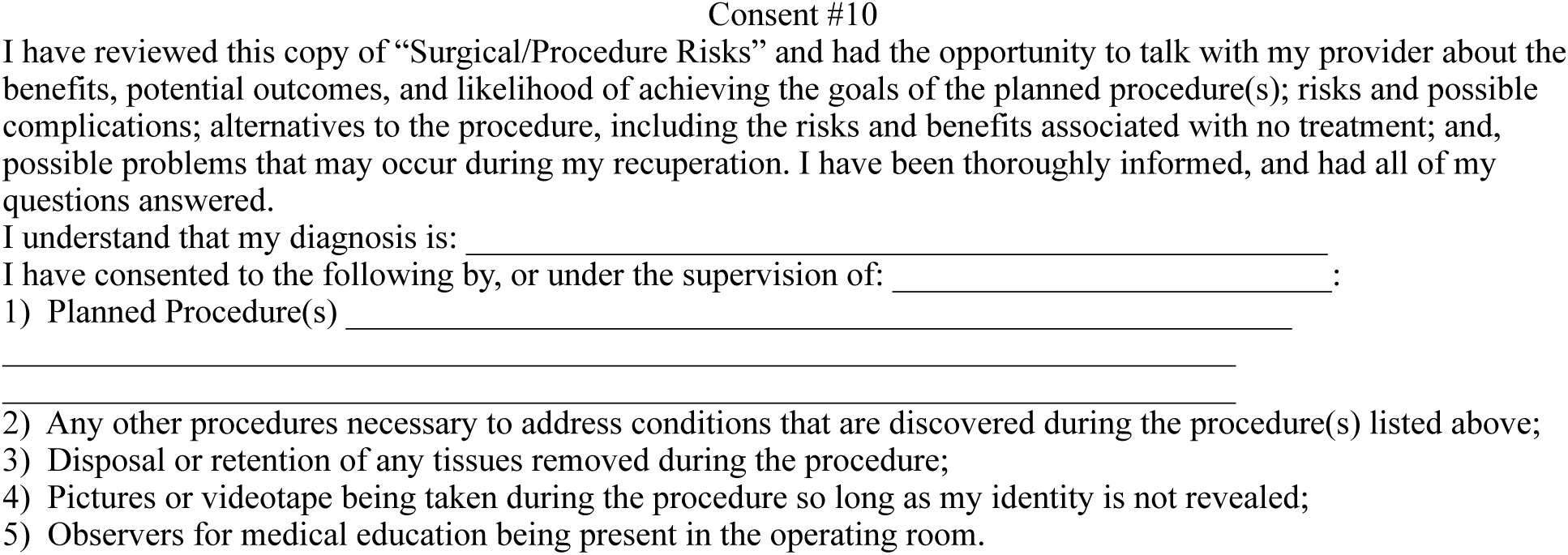
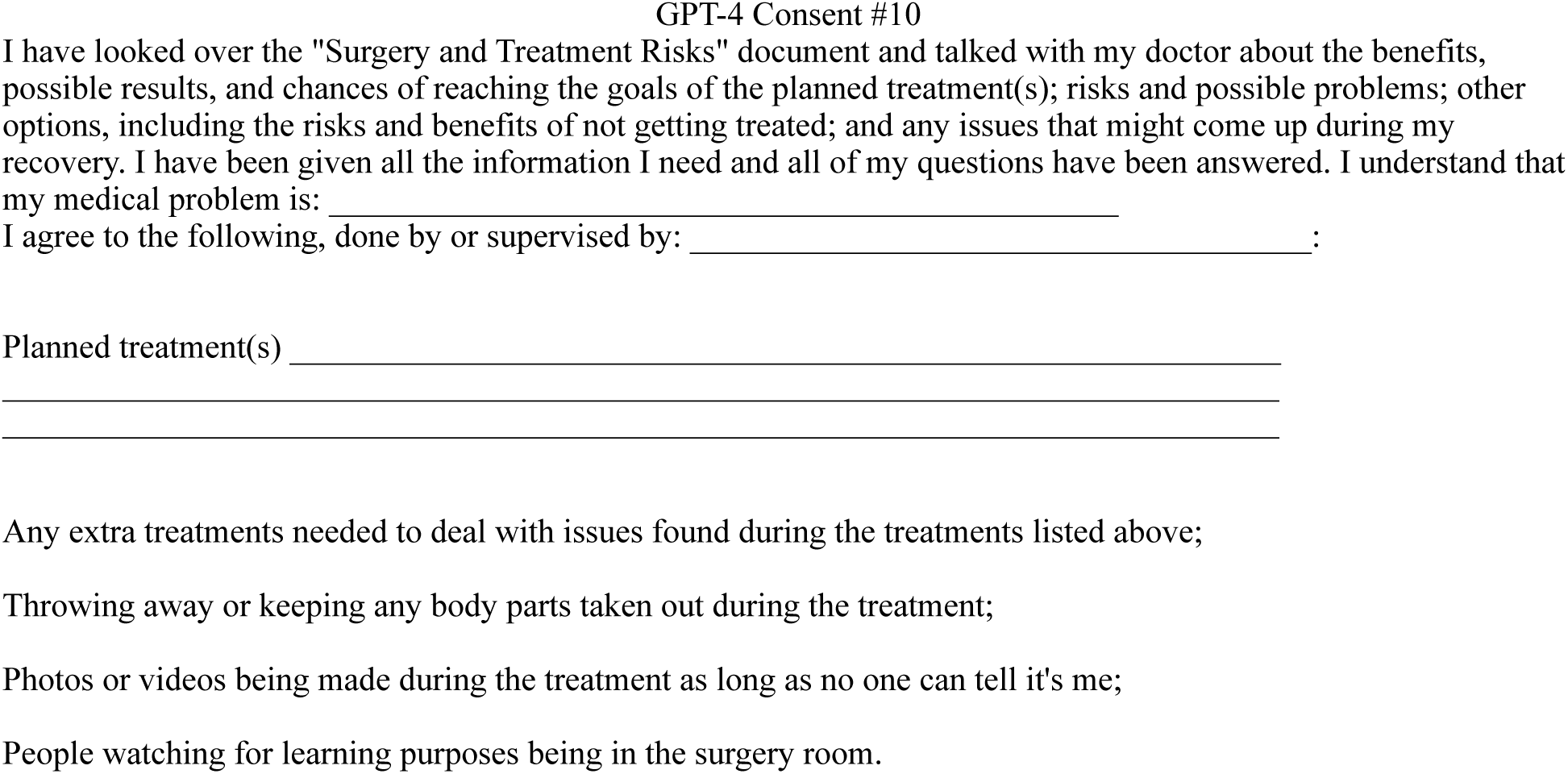
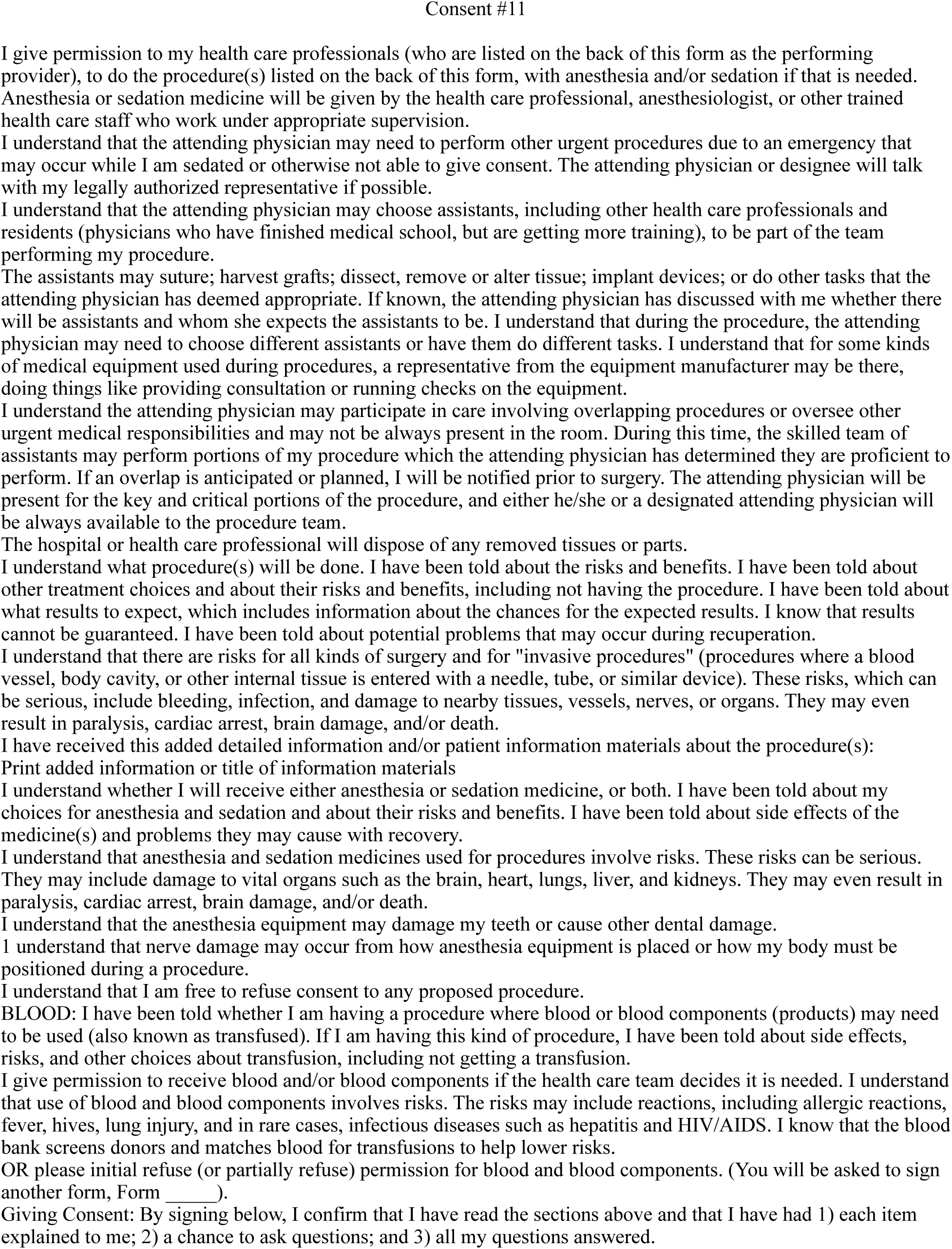
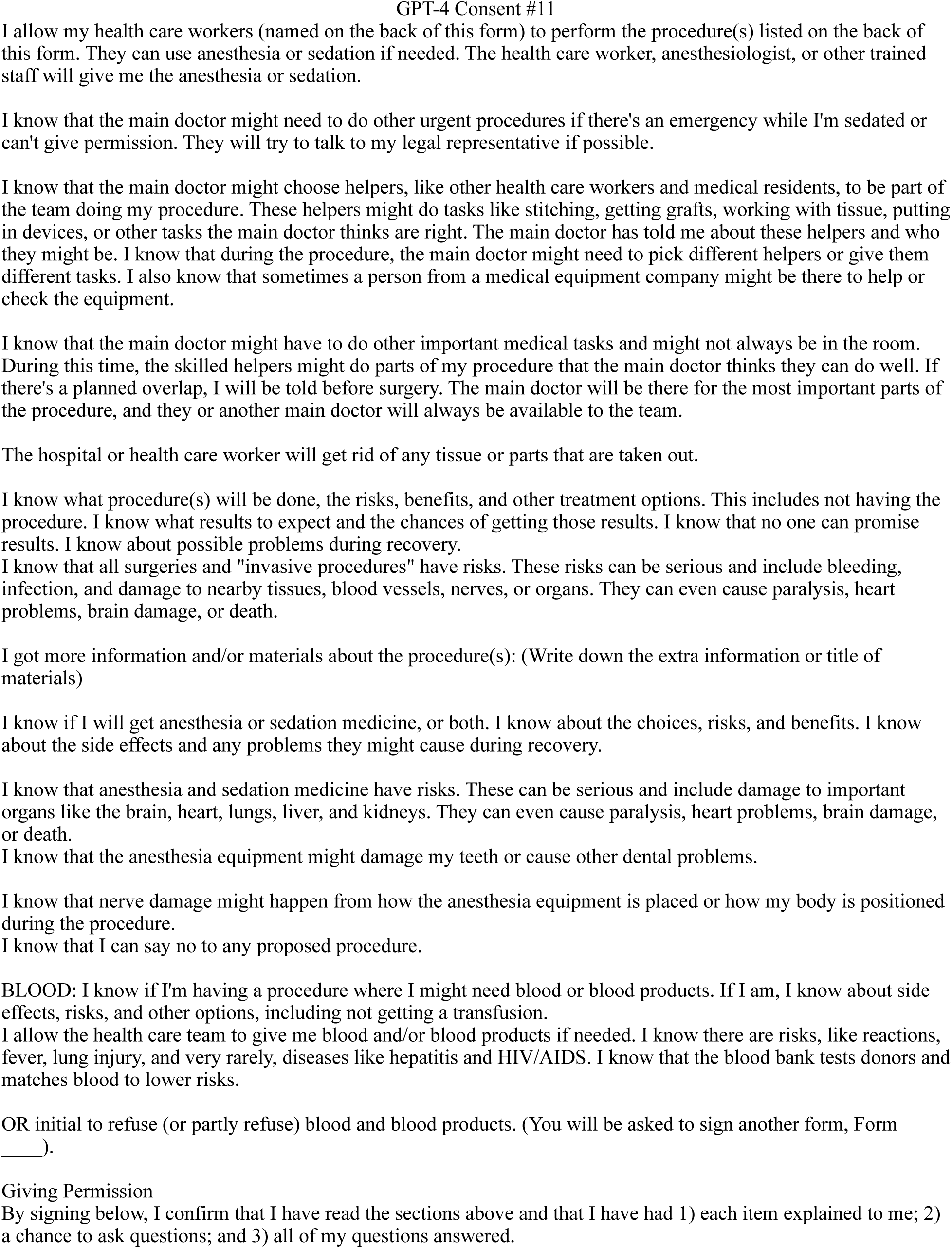
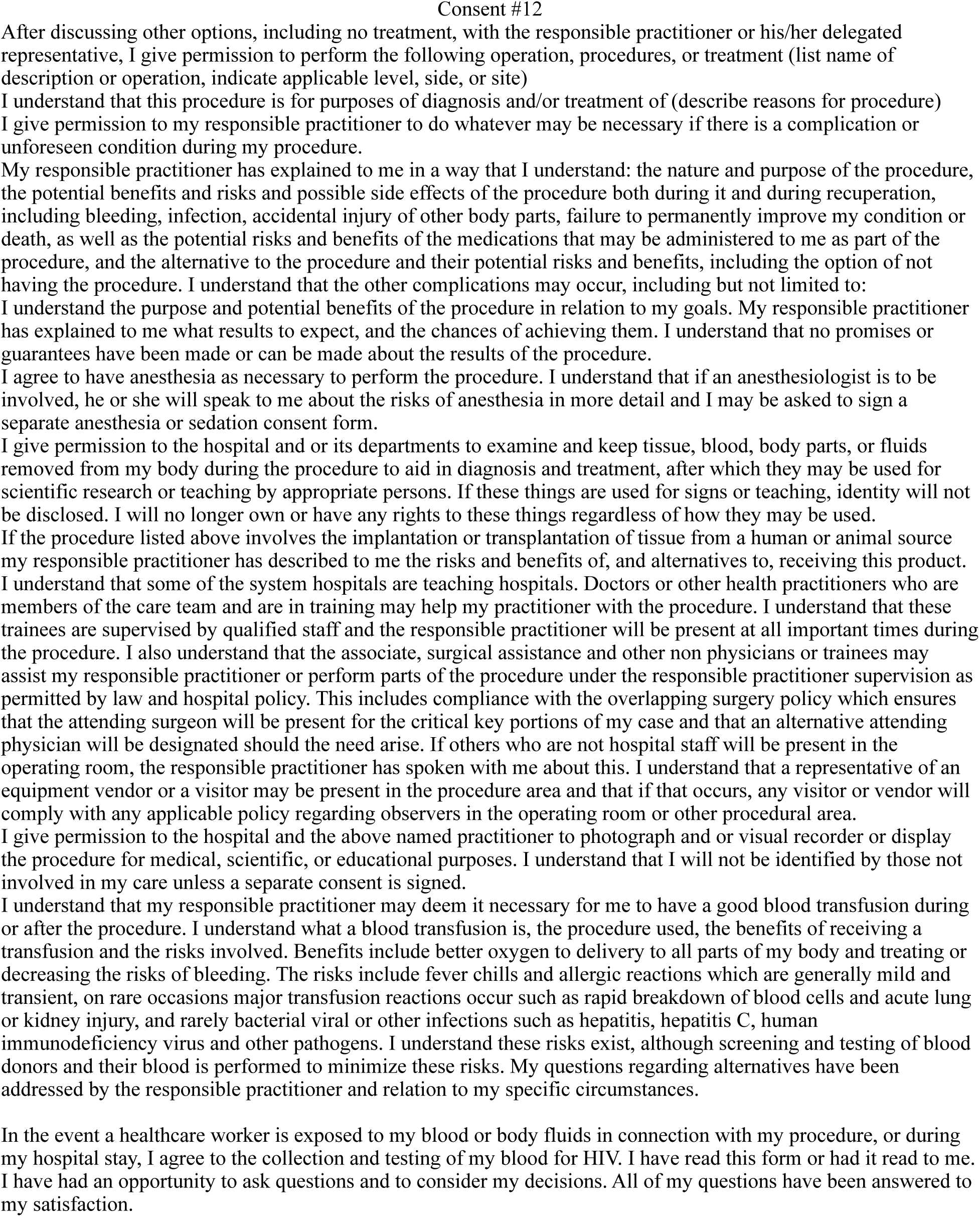
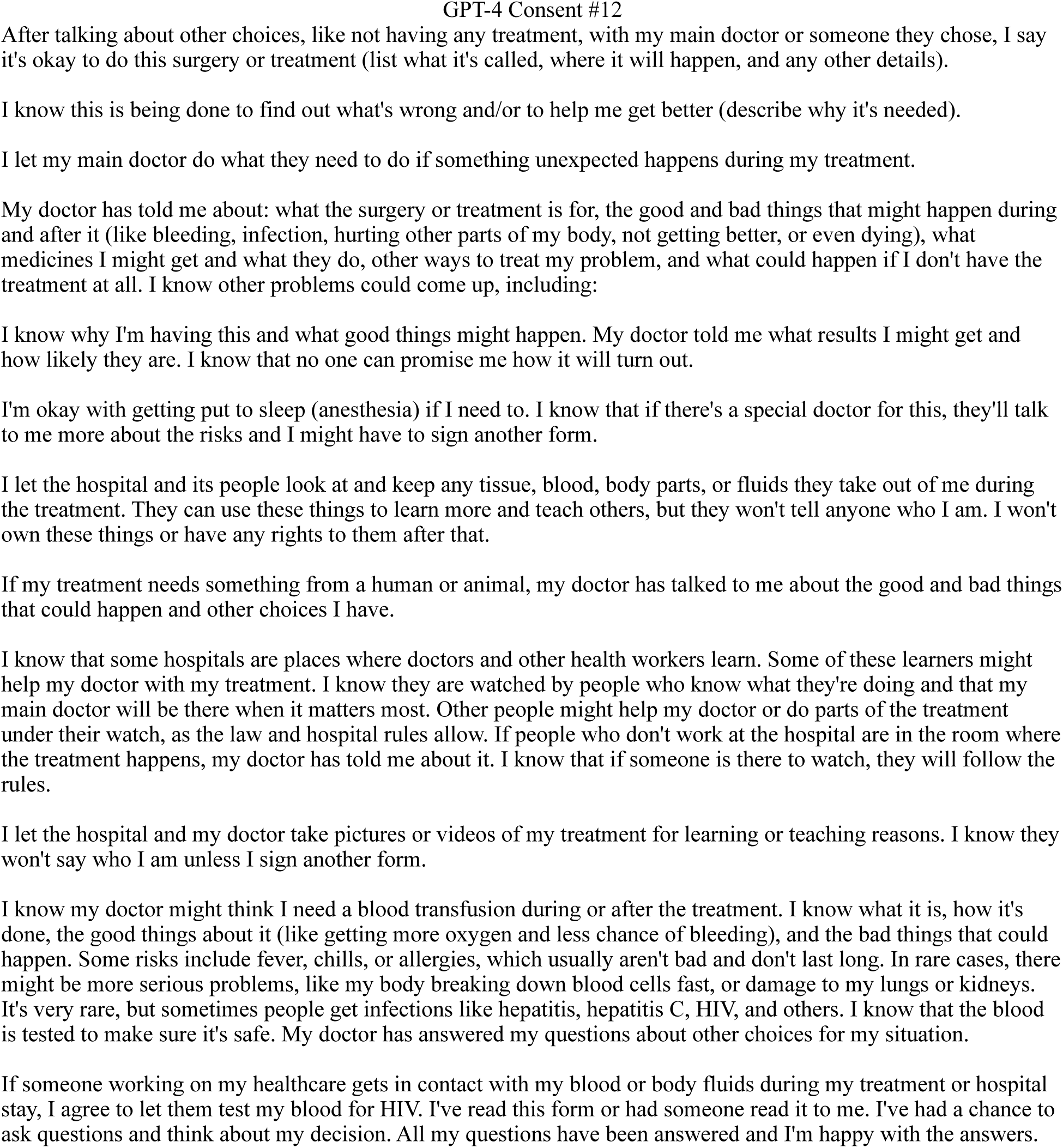
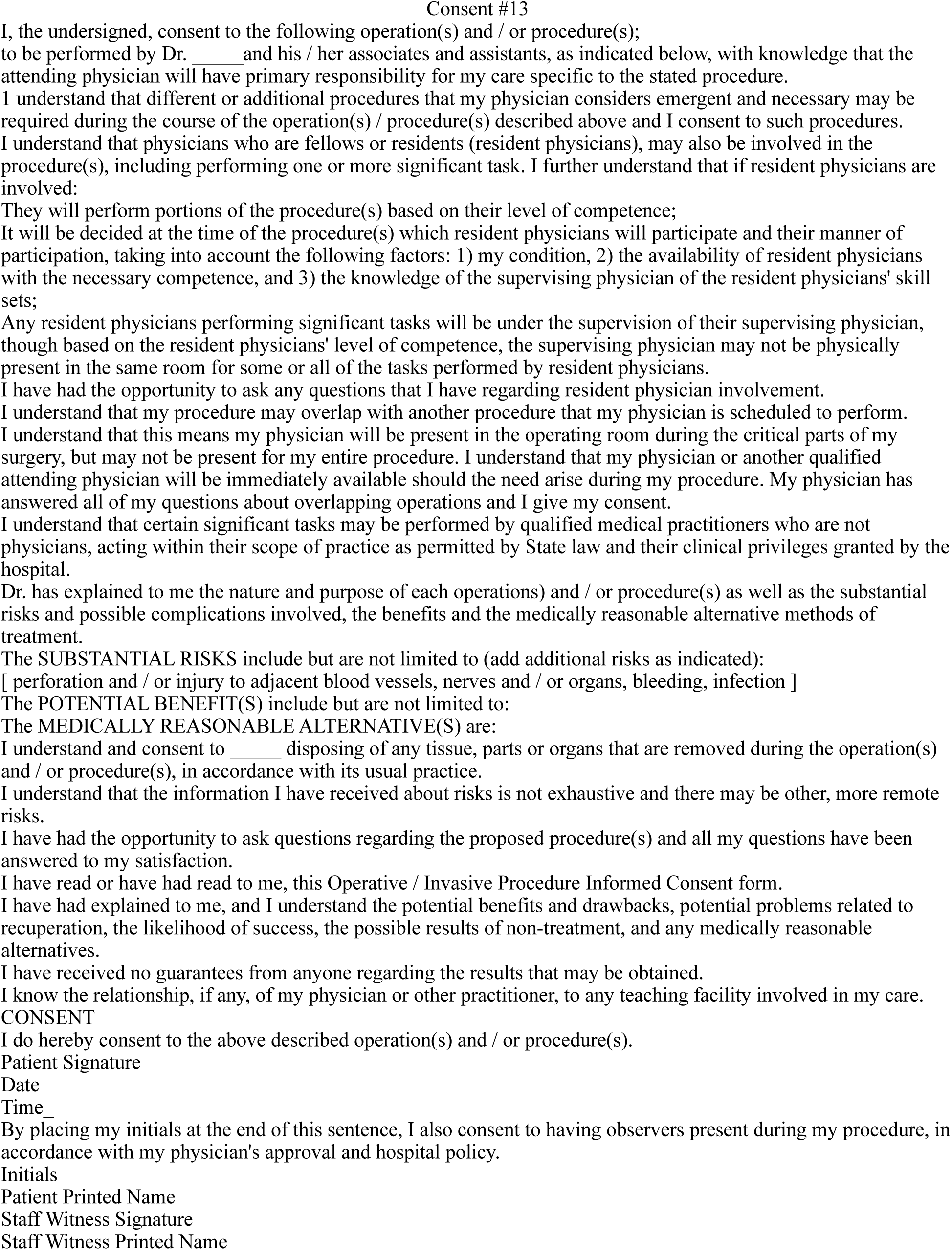
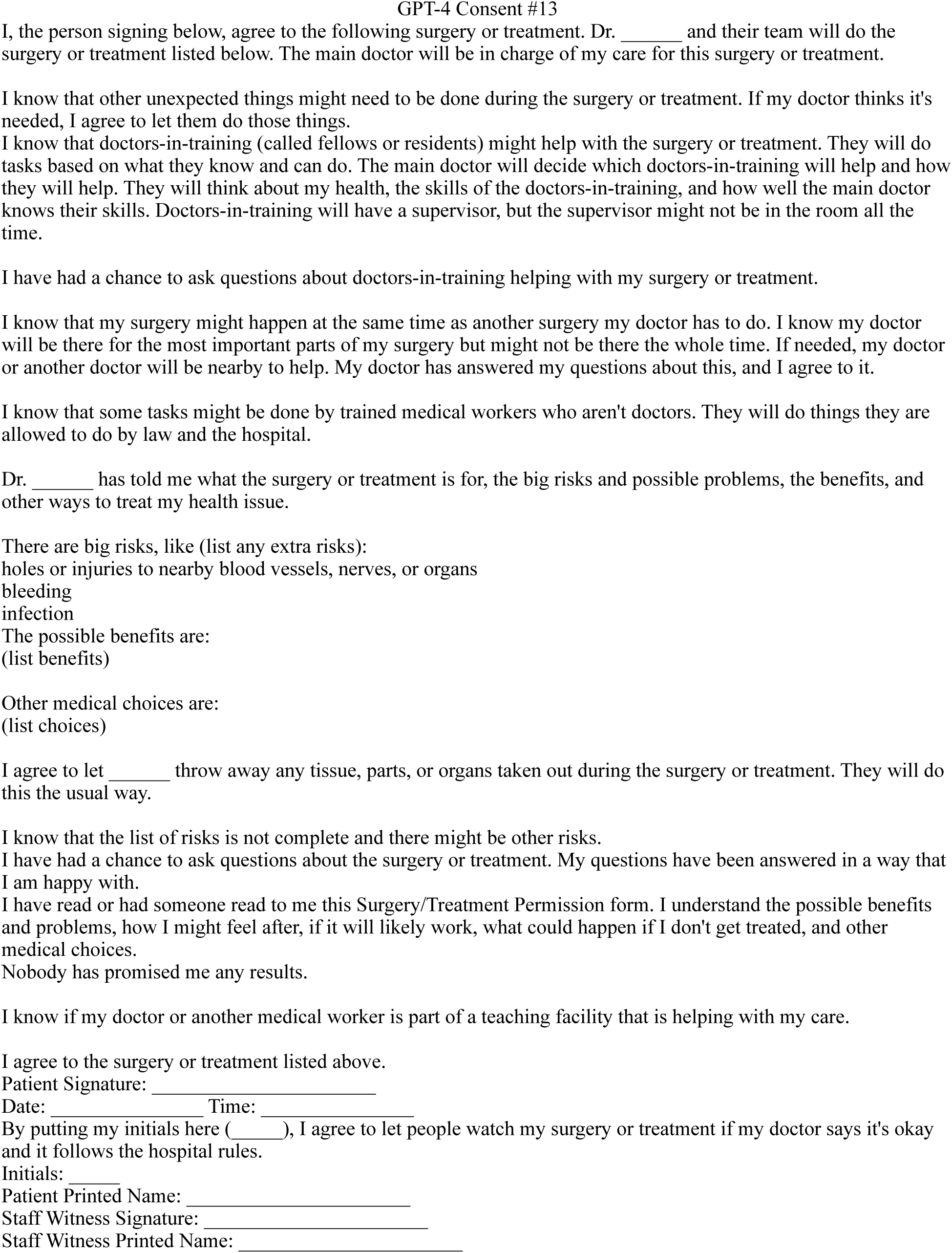
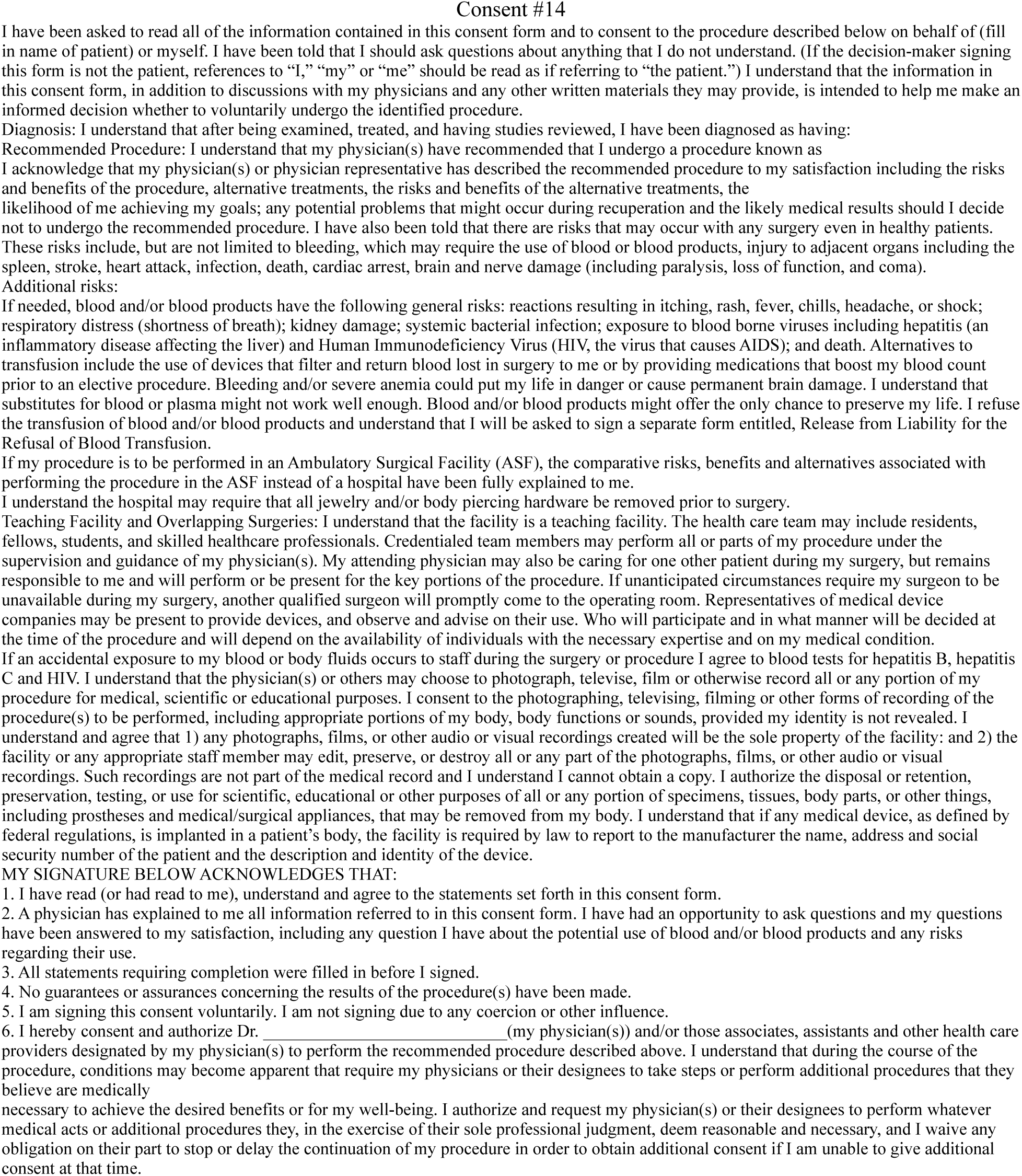
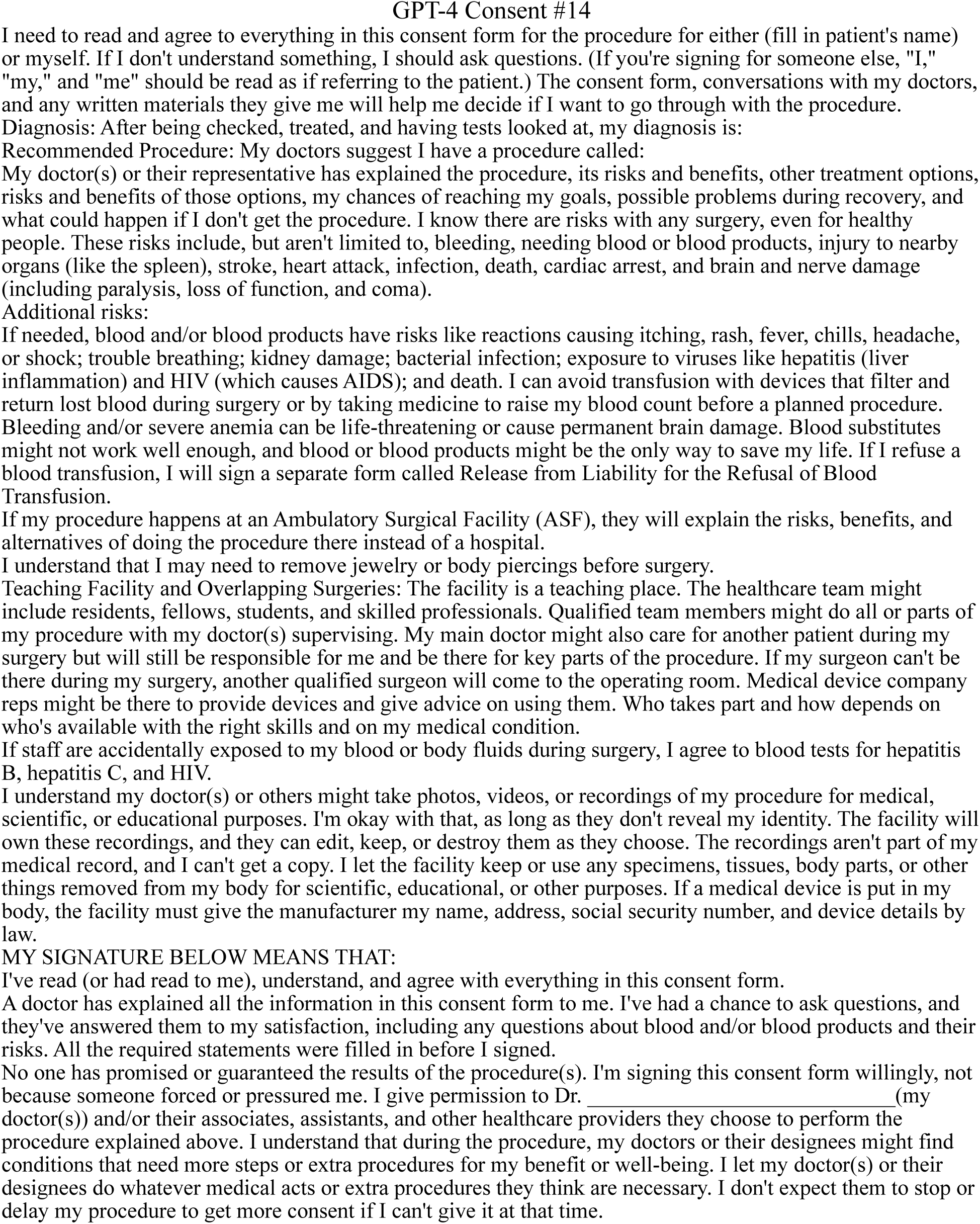
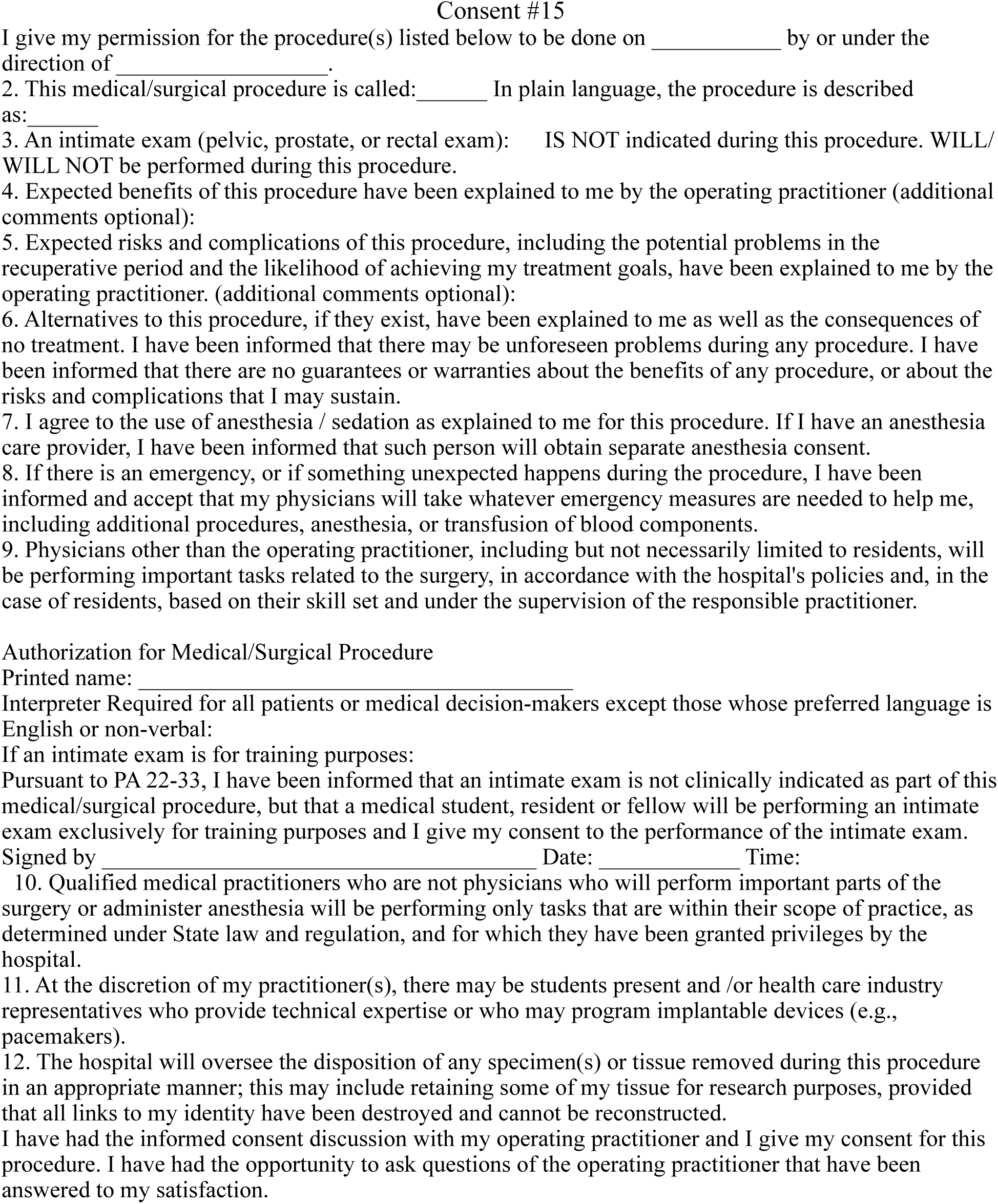
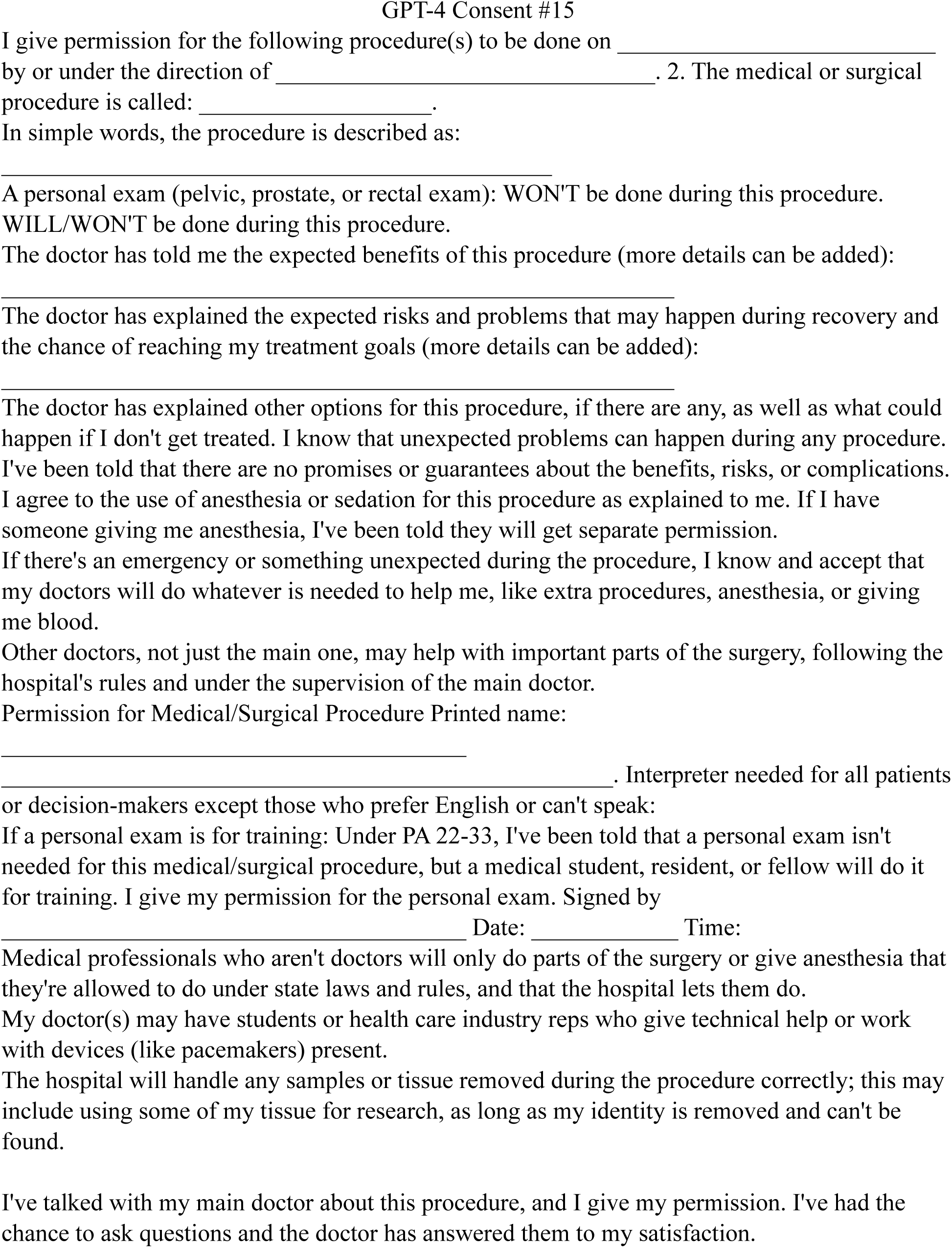
Generic Consent Forms at 15 Academic Medical Centers. Copies of the 15 generic consent forms prior to and after undergoing LLM-facilitated simplification.

